# Distress above diagnostic constraints: transdiagnostic psychological and somatic symptom patterns in young adults

**DOI:** 10.64898/2026.03.25.26349193

**Authors:** Ann-Kathrin Schwientek, Julia Braun, Andreas M. Baumer, Viktoriia Yasenok, Viktoriia Petrashenko, Marco Kaufmann, Anja Frei, Seraina Rüegger, Tala Ballouz, Andrii Loboda, Vladyslav Smiianov, Susi Kriemler, Viktor von Wyl, Susanne Walitza, Andriana Kostenko, Stefan Büchi, Milo A. Puhan

## Abstract

**Background:** Somatic and psychological symptoms like depression, anxiety, and trauma-related stress often co-occur, especially in young adults, a group facing major life transitions and increased vulnerability. These overlapping symptoms pose diagnostic challenges that traditional disorder-specific models capture poorly. Transdiagnostic and dimensional approaches may offer a more meaningful framework. However, population-based data on symptom patterns in young adults remains sparse. This study investigated the patterns of psychological and somatic symptoms among young adults from Switzerland and compares these results to findings from populations with different stress exposure histories: Ukrainians who fled to Switzerland, and Ukrainians living in different regions in Ukraine during the war.

**Methods:** We analyzed cross-sectional baseline data collected in spring 2024 as part of the Mental Health Assessment of the Population (MAP) studies, where we enrolled randomly selected young adults aged 18–24 from Switzerland, Ukrainian refugees in Switzerland, and Ukrainians residing in regions with different degrees of proximity to active war zones. We assessed somatic (PHQ-15) and psychological symptoms (PHQ-9, GAD-7, PCL-5) and explored symptom patterns using descriptive statistics, correlations, and k-means clustering.

**Results:** Psychological symptom severity showed highly consistent moderate-to-strong correlations with somatic symptoms (range: 0.53-0.69), across all young adult subgroups and disorders. Rather than identifying disorder-specific patterns, symptoms clustered by overall symptom severity, emerging in three clusters: (1) high symptom burden, (2) moderate symptom burden, and (3) low symptom burden clusters with elevated somatic, depressive, anxiety, and PTSD symptoms. The cluster structure was remarkably stable across Swiss, Ukrainian, and refugee subsamples, despite markedly different stress exposure histories.

**Conclusion:** Our results support a symptom-based, dimensional approach to understanding mental health in young adults and to better capture the complexity and co-occurrence of psychological and somatic symptoms in this age group. These findings further suggest that prevention and early detection strategies should more systematically integrate both psychological and somatic symptomatology.

## Background

Psychological symptoms seen in depression, anxiety, and trauma-related stress frequently co-occur with somatic symptoms [1–5], posing complex diagnostic and therapeutic challenges for health care professionals [3, 6]. Young adulthood is a developmental stage characterized by major life transitions, physiological and psychosocial changes, and increased stress exposure, where vulnerability to psychological and somatic symptoms is heightened and may carry long-term implications for health and functioning [7–12]. International data indicate a high prevalence of these symptoms among adolescents and young adults [13–20]. Such symptoms, even mild complaints, can impair daily functioning if they persist and are associated with poorer health status and increased healthcare utilization, regardless of underlying medical pathology [1, 5, 7, 21–23]. Young adults with mental health problems and chronic conditions face substantial difficulties entering the labor market and remaining in employment, with far-reaching economic consequences [24, 25]. In Switzerland, despite an overall decline in disability pension inflows, disability pension rates among young adults have not decreased, highlighting the need for early, targeted interventions [25].

The association and overlaps of somatic symptoms and mental health conditions complicates differential diagnosis and treatment planning [4, 26, 27]. Clear categorial diagnostic boundaries are poorly supported by empirical evidence [26, 27]. Criticism of categorical diagnostic systems has a long history, with transdiagnostic and dimensional models having been discussed and developed for decades [28, 29]. Despite this, categorical approaches have remained the standard in clinical practice and treatment decisions [29]. However, recent shifts in the DSM-5 and ICD-11 better reflect a symptom- and function-based approach [30, 31]. Psychological research has moved toward dimensional and transdiagnostic models that use continuous indicators of psychopathology across diagnostic categories and emphasize shared underlying constructs [32]. These transdiagnostic approaches, such as the Hierarchical Taxonomy of Psychopathology (HiTOP) model, the Research Domain Criteria (RDoC) matrix, and the Network Theory (NT), better reflect the complexity, dimensionality and comorbidity encountered in clinical practice [32–40].

Despite this growing awareness of health challenges among young adults, existing studies have often focused on singular medical or psychiatric conditions rather than exploring patterns of co-occurring symptoms in the general population [5, 13, 14, 41–43]. As a result, the heterogeneity and complexity of symptom presentations in this age group remain insufficiently understood, contributing to diagnostic delays and fragmented care [44]. Previous research shows a high somatic symptom burden among youth with mental disorders, with somatic symptoms reported in up to 70% of adolescents with anxiety or depression [8]. Similar findings have been observed in clinical populations in Norway and Germany [18, 21, 45–47]. Current evidence highlights the urgent need for better understanding the interaction of psychological and somatic symptoms [16, 20, 23] in order to inform early detection, diagnostic approaches, and integrative care strategies for young adults [5, 18]. However, population-level data on transdiagnostic symptom patterns in young adulthood remain sparse. Therefore, this study aimed to investigate the co-occurrence and clustering patterns of somatic and psychological symptoms, in a population-based sample of young adults and further extend previous work by incorporating populations with different life experiences and stress exposures.

## Methods

### Study design and setting

We used baseline data from the MAP studies [48], which are prospective, population-based, observational cohort studies encompassing three populations: The Zurich, Switzerland, general population (MAP-Z (ZH)), refugees from Ukraine residing in the canton of Zurich (MAP-Z (UA)), and the general population of Ukraine (MAP-U). A detailed study protocol [48] and the first baseline results have been published [20]. For detailed information and the full rationale of the sample size calculation, we refer to the MAP protocol and baseline paper [20, 48].

In brief, the MAP studies aim to enable public mental health surveillance through an online monitoring of psychological symptoms among the general population and relevant subgroups. To better reflect the different levels of war-related stress exposure within Ukraine (MAP-U), we focused on three regional clusters formed from 10 selected oblasts across different parts of the country, each experiencing varying degrees of impact from the ongoing war. At the time of recruitment, the North-West cluster (NW), including Rivne, Lviv, and Chernivtsi oblasts, was located far from the active war zone and experienced comparatively low security risks. The Central cluster (CE), including Zhytomyr, Kyiv, and Dnipropetrovsk oblasts, was closer to the border to Russia and experienced moderate security risks. The South-East cluster (SE), including Sumy, Kharkiv, Mykolaiv, and Kherson oblasts, was situated near active war zones, and faced high security risks (**Figure S1).**

The analysis described in this paper focuses on the subgroup of young adults, aged 18-24 years. Accordingly, we describe only the methods and results relevant to this study population.

### Eligibility criteria and recruitment

The study enrolled participants aged ≥ 18 years, who were capable of completing an online survey and provided informed consent. For MAP-Z (ZH) we included individuals residing in the Canton of Zurich, holding either Swiss citizenship or residence permit B (time-limited residence and work permit for foreign nationals planning to stay in Switzerland for more than a year) or C (settlement permit that grants permanent residency rights). Participants in MAP-Z (UA) were Ukrainian nationals in the canton of Zurich, registered with protection status S (temporary residency permit granted to individuals fleeing conflict until returning home is considered safe). For MAP-U, eligible participants were living in one of the ten oblasts of Ukraine described above, who provided informed consent and were able to complete the online survey. Repeated unsuccessful contact attempts lead to an exclusion of the study. Recruitment was conducted between 25 March and 9 August 2024.

We employed age-stratified random sampling based on population registries and made substantial efforts to enhance participation rates. MAP-Z (ZH) and MAP-Z (UA) potential participants were randomly selected by the Swiss Federal Statistical Office and initial invitations were sent by postal mail [49, 50]. MAP-U potential participants were randomly selected by the Center for Social Research at Sumy State University based on population registries of the oblasts. Selected persons were contacted via email, including two reminders. If there was no response, the interviewers from our local teams in Ukraine approached them via Telegram (until May 2024 when Ukrainians were recommended to stop using it), Viber (from May 2024) or phone calls.

All recruited participants received information about the study, an invitation to participate and a personalized link and access code to a secure digital study platform (REDCap) [51, 52], where they received further information. Those interested provided informed consent to participate in the study electronically via REDCap and were forwarded to the baseline assessment. Follow-up assessment occurs every three months for at least two years, with invitations sent by email, Telegram or Viber.

Eligible for the inclusion in the present analysis were all participants who have completed the baseline questionnaire.

### Data collection and instruments

We collected sociodemographic data and assessed mental health outcomes, including symptoms of PTSD (Posttraumatic Stress Disorder Checklist, PCL-5), anxiety (Generalised Anxiety Disorder 7-item scale, GAD-7), depression (Patient Health Questionnaire-9, PHQ-9), as well as somatic distress (Patient Health Questionnaire-15, PHQ-15). In addition, we assessed preexisting chronic physical and mental health conditions of young adults. Instruments were selected based on psychometric quality, established cut-offs, digital feasibility, population relevance, prior use in Ukraine, and alignment with the Global Burden of Disease (GBD) standards [53]. Only participants with complete sets of all four surveys were included in our analyses. In our study population, internal consistency reliability was high: PHQ-9 (α = 0.86), GAD-7 (α = 0.90), PCL-5 (α = 0.94), and PHQ-15 (α = 0.82). More details about the instruments can be found in the MAP protocol and baseline paper [20, 48]. Detailed information on the cut-points for each instrument, as well as their sensitivity and specificity, is provided in the supplementary material.

### Statistical analysis

Descriptive analyses were conducted using means with standard deviations (SD), medians and interquartile ranges (IQR) for continuous data and sum scores, and absolute numbers with percentages for categorical data. Correlations between individual PHQ-15 items and PHQ-9, GAD-7 and PCL-5 items were further explored using Spearman’s correlation coefficient.

To identify symptom burden patterns, we performed k-means clustering using PHQ-15, PHQ-9, GAD-7 and PCL-5 items [54]. The number of clusters was chosen based on the gap statistic [55]. For MAP-U we performed all data analyses stratified by the three regions in Ukraine (North-West, Central, South-East) comparing the results across the different subsamples for different war-related exposure.

Sensitivity analyses were conducted to assess the influence of missing values in any of the items on the clustering: To evaluate the maximum range of missing values, we performed single imputation of the minimal and maximal value of the respective items and repeated the cluster analysis. We conducted all analyses in R 4.5.2.

## Results

### Sample characteristics

Information about the recruitment process detailed as a Consolidated Standards of Reporting Trial diagrams as well as participation rates can be found in our published baseline paper [20]. We analyzed data from all participants who have completed the baseline assessment, which lead to n=142 participants in MAP-Z (ZH), n=102 in MAP-Z (UA) and n=1176 in the MAP-U subgroup. As only patients with no missing data in any of the items could be included in the cluster analysis, these numbers were reduced to n=127 participants in the MAP-Z (ZH), n=91 in the MAP-Z (UA) and n=963 in the MAP-U subgroup.

Sociodemographic characteristics and existing chronic health conditions of young adults among the different subgroups are detailed in **Table 1**. A sex imbalance with more women (57.5% to 77.4%) was observed in all samples. Across all study arms about 22.0% to 40.6% of the young adults reported preexisting chronic health conditions, including physical (cardiovascular, nervous, stroke, respiratory, oncological, diabetes, digestive, genitourinary, musculoskeletal, allergic conditions) and mental health conditions. While the overall prevalence in MAP-Z (ZH) and MAP-Z (UA) was comparatively lower, the proportion of psychiatric conditions was much higher at 15.0% and 12.1% in comparison to MAP-U (NW) (4.7%), MAP-U (CE) (7.4%) and MAP-U (SE) (6.7%).

**Table 1.**
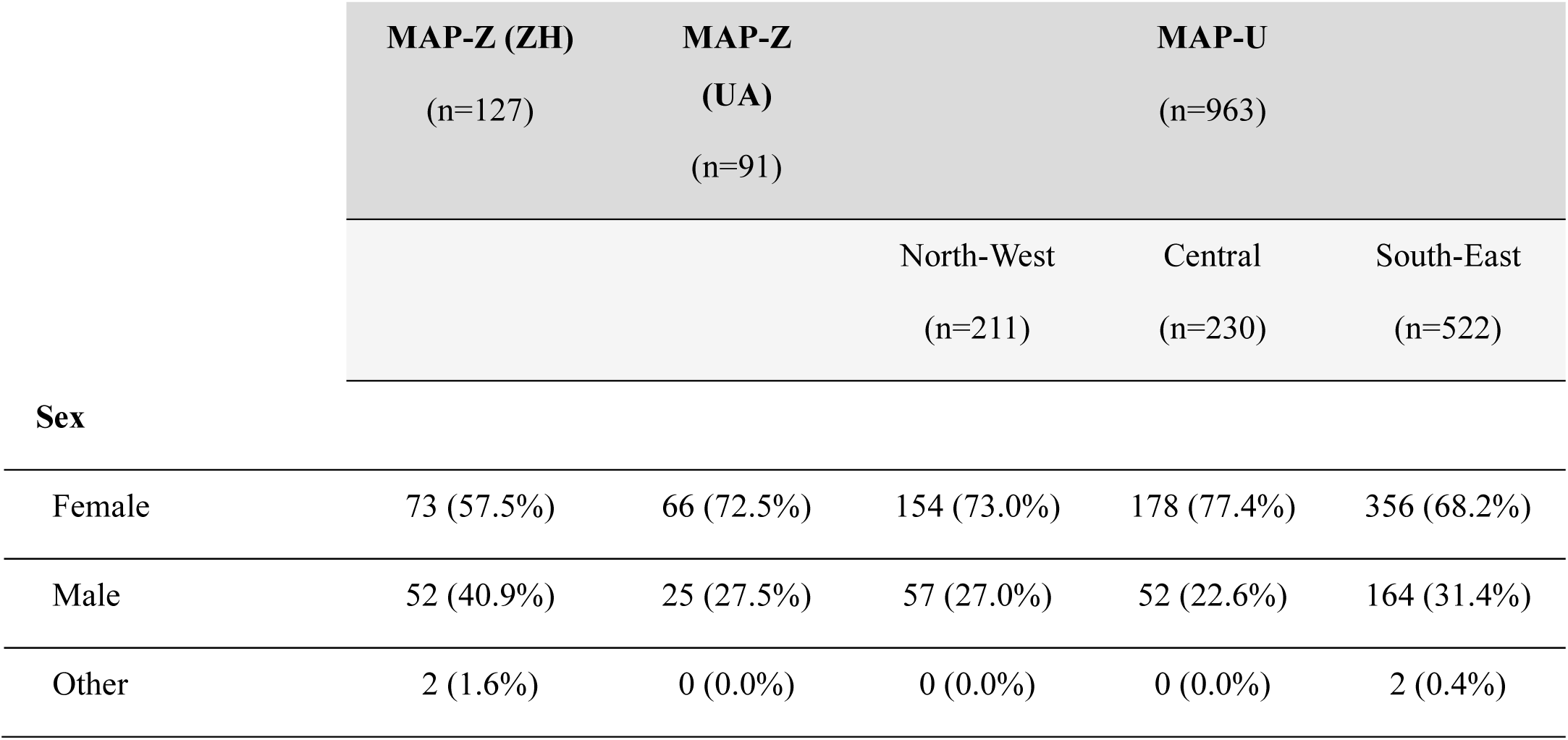
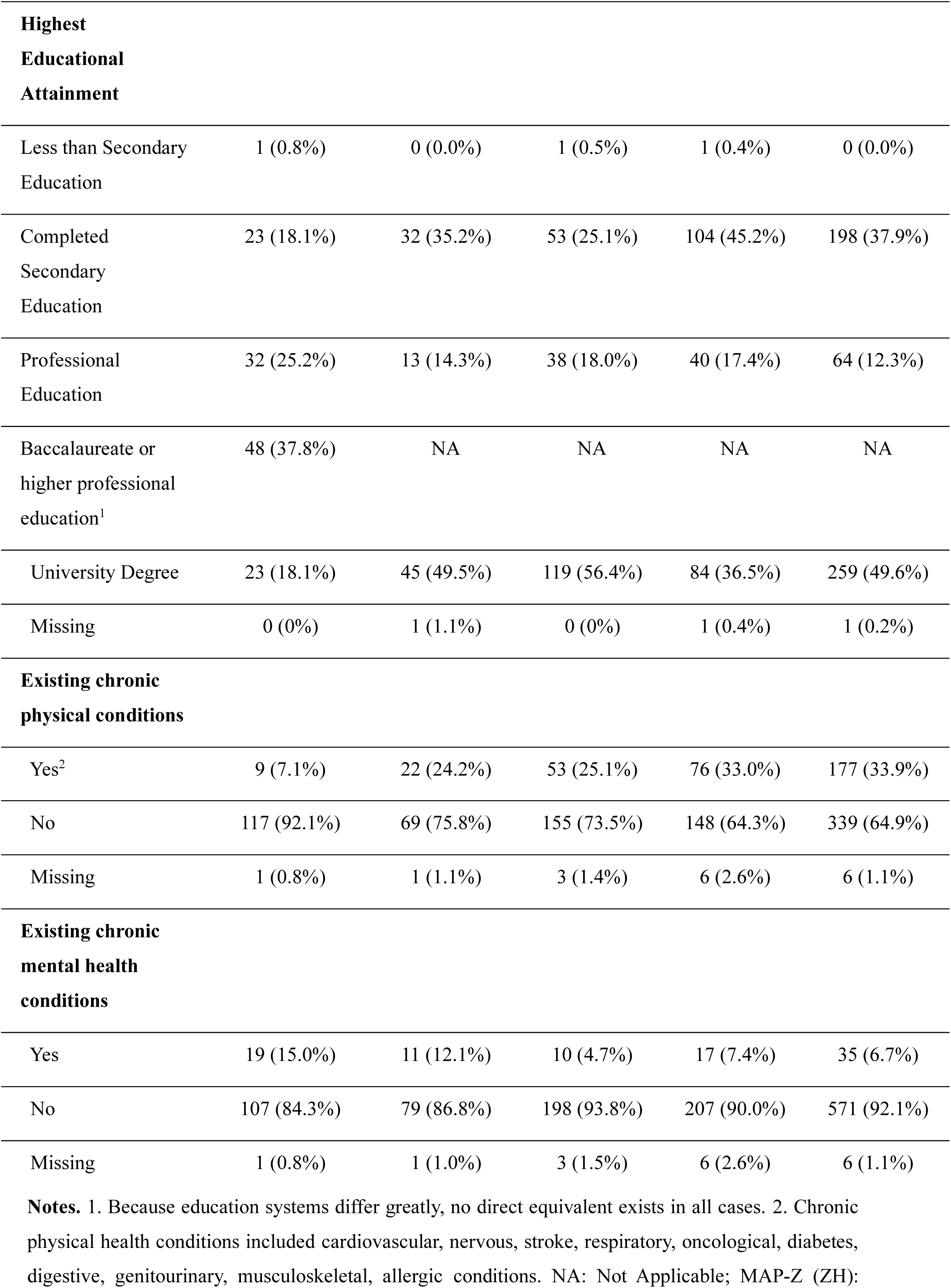
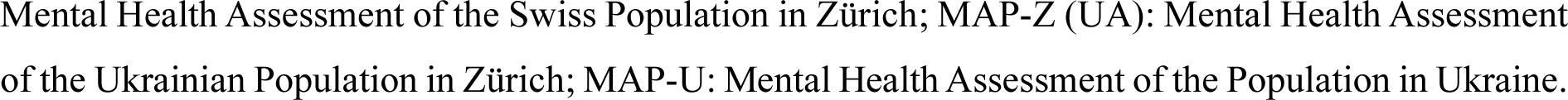
Demographic characteristics and health conditions of MAP-Z (ZH), MAP-Z (UA), MAP-U age grouped 18-24 years old (in total n = 1181 participants).

|  | MAP-Z (ZH)<br>(n=127) | MAP-Z<br>(UA)<br>(n=91) | MAP-U<br>(n=963) |  |  |
| --- | --- | --- | --- | --- | --- |
|  |  |  | North-West<br>(n=211) | Central<br>(n=230) | South-East<br>(n=522) |
| **Sex** |  |  |  |  |  |
| Female | 73 (57.5%) | 66 (72.5%) | 154 (73.0%) | 178 (77.4%) | 356 (68.2%) |
| Male | 52 (40.9%) | 25 (27.5%) | 57 (27.0%) | 52 (22.6%) | 164 (31.4%) |
| Other | 2 (1.6%) | 0 (0.0%) | 0 (0.0%) | 0 (0.0%) | 2 (0.4%) |
| **Highest Educational Attainment** |  |  |  |  |  |
| Less than Secondary Education | 1 (0.8%) | 0 (0.0%) | 1 (0.5%) | 1 (0.4%) | 0 (0.0%) |
| Completed Secondary Education | 23 (18.1%) | 32 (35.2%) | 53 (25.1%) | 104 (45.2%) | 198 (37.9%) |
| Professional Education | 32 (25.2%) | 13 (14.3%) | 38 (18.0%) | 40 (17.4%) | 64 (12.3%) |
| Baccalaureate or higher professional education<sup>1</sup> | 48 (37.8%) | NA | NA | NA | NA |
| University Degree | 23 (18.1%) | 45 (49.5%) | 119 (56.4%) | 84 (36.5%) | 259 (49.6%) |
| Missing | 0 (0%) | 1 (1.1%) | 0 (0%) | 1 (0.4%) | 1 (0.2%) |
| **Existing chronic physical conditions** |  |  |  |  |  |
| Yes<sup>2</sup> | 9 (7.1%) | 22 (24.2%) | 53 (25.1%) | 76 (33.0%) | 177 (33.9%) |
| No | 117 (92.1%) | 69 (75.8%) | 155 (73.5%) | 148 (64.3%) | 339 (64.9%) |
| Missing | 1 (0.8%) | 1 (1.1%) | 3 (1.4%) | 6 (2.6%) | 6 (1.1%) |
| **Existing chronic mental health conditions** |  |  |  |  |  |
| Yes | 19 (15.0%) | 11 (12.1%) | 10 (4.7%) | 17 (7.4%) | 35 (6.7%) |
| No | 107 (84.3%) | 79 (86.8%) | 198 (93.8%) | 207 (90.0%) | 571 (92.1%) |
| Missing | 1 (0.8%) | 1 (1.0%) | 3 (1.5%) | 6 (2.6%) | 6 (1.1%) |

**Notes.** 1. Because education systems differ greatly, no direct equivalent exists in all cases. 2. Chronic physical health conditions included cardiovascular, nervous, stroke, respiratory, oncological, diabetes, digestive, genitourinary, musculoskeletal, allergic conditions. NA: Not Applicable; MAP-Z (ZH):
Mental Health Assessment of the Swiss Population in Zürich; MAP-Z (UA): Mental Health Assessment of the Ukrainian Population in Zürich; MAP-U: Mental Health Assessment of the Population in Ukraine.

In addition, **Table S1** in the supplementary material summarizes direct and indirect (through relatives) exposure to war-related events in MAP-Z (UA) and MAP-U providing background information on war-related stress exposures.

Cross tables of psychological and somatic symptoms using the screening tool cut-offs detailed in the methods section are presented also in the supplementary material (**Table S2 – Table S6**). The results indicate a moderate to high comorbidity for clinically relevant somatic symptoms, depression, anxiety, and PTSD among all study arms (MAP-Z (ZH), MAP-Z (UA), MAP-U (NW), MAP-U (CE) and MAP-U (SE)).

### Main results

We first give an overview of the symptom burden across all subgroups of young adults detailed in **Figure 1** for somatic symptoms, depression, anxiety and PTSD symptoms. Young Ukrainian adults living in Ukraine and Switzerland showed a higher overall burden of somatic symptoms, anxiety as well as post-traumatic stress than the general population of young adults in Switzerland which showed a low to moderate symptom burden. Within the Ukrainian subsample MAP-U, MAP-U (SE) and MAP-U (CE) showed a higher symptom burden across all instruments in comparison to MAP-U (NW). Also, Ukrainians living in Switzerland with protection status S (MAP-Z (UA)) reported higher symptom burdens in comparison to MAP-U (NW), comparable to those of MAP-U (SE, CE). Detailed results for all sum scores across instruments and subgroups can be found in the supplementary material (**Table S7)**.

**Figure 1.**
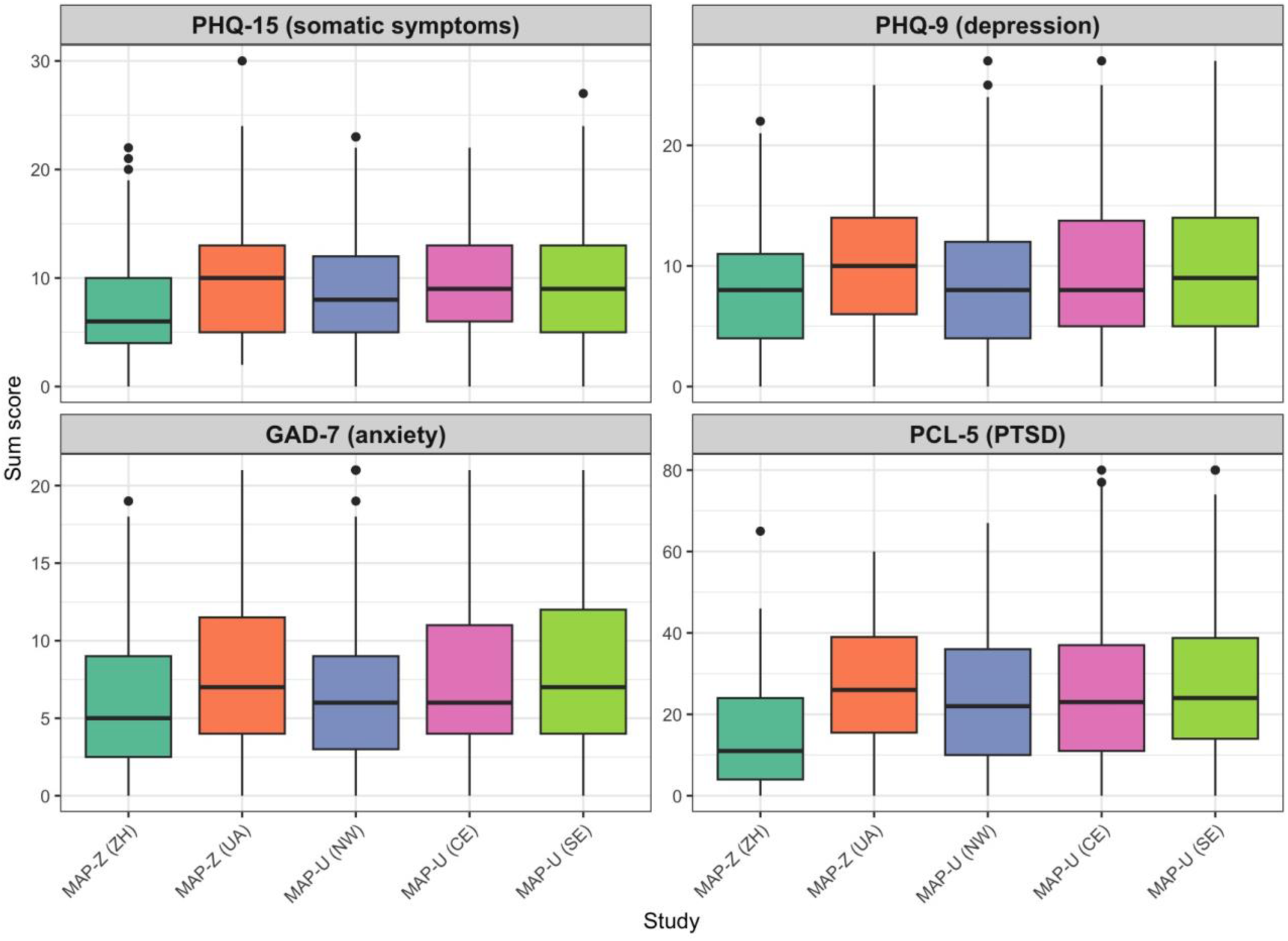
Boxplots of PHQ-15, PHQ-9, GAD-7 and PCL-5 sum scores by MAP study arms MAP-U (NW), MAP-U (SE), MAP-U- (CE), MAP-Z (UA) and MAP-Z (ZH) in 18-24 years old. **Notes.** MAP-Z (ZH): Mental Health Assessment of the Swiss Population in Zürich; MAP-Z (UA): Mental Health Assessment of the Ukrainian Population in Zürich; MAP-U (NW): Mental Health Assessment of the Population in Ukraine (North-West); MAP-U (CE): Mental Health Assessment of the Population in Ukraine (Central); MAP-U (SW): Mental Health Assessment of the Population in Ukraine (South-East); PHQ-15: Patient Health Questionaire-15; PHQ-9: GAD-7: Generalized Anxiety Disorder-7; Patient Health Questionaire-9; PCL-5: PTSD Checklist for DSM-5.

In the supplementary material, we provide an overview of the frequency of individual items from the PHQ-15 reported by young adults in MAP-Z (ZH), MAP-Z (UA), and MAP-U (**Figures S2–S4**). Among all study arms the most frequently reported somatic complaints were fatigue, sleep difficulties, back pain and headache. For the MAP-Z (UA) and MAP-U also the symptoms heart racing and shortness of breath were more frequently reported.

The correlations between somatic and psychological symptoms are illustrated in scatterplots shown in **Figure 2**. PHQ-15 sum scores were positively correlated with depressive (PHQ-9), anxiety (GAD-7), and PTSD (PCL-5) symptom scores. Correlations across all instruments and across all subgroups were very similar, ranging from r = 0.53 to r = 0.69 (moderate to strong correlations).

**Figure 2.**
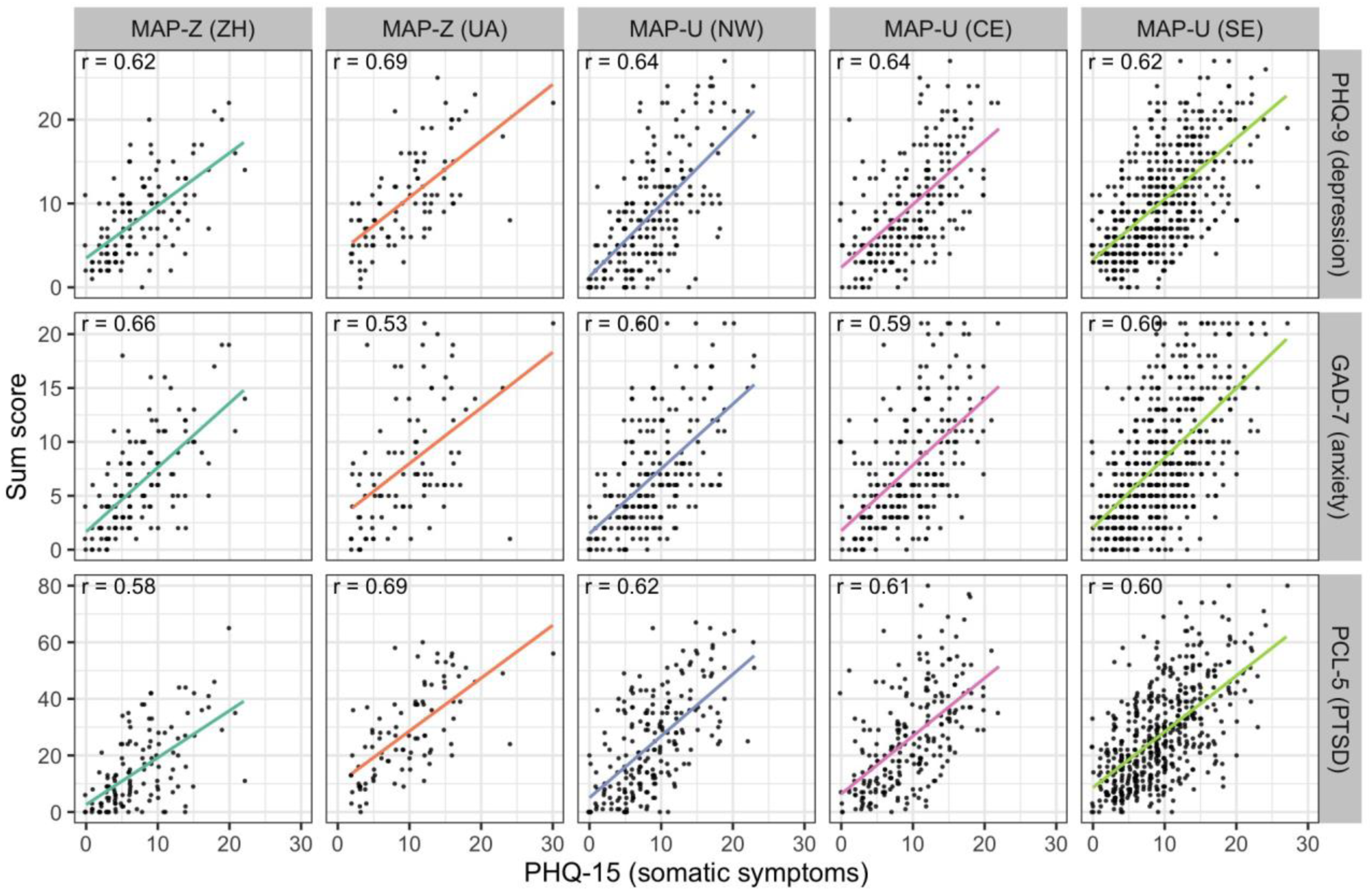
Scatter Plot with regression lines describing the correlation between PHQ-15 with PHQ-9, GAD-7 and PCL-5 sum scores in MAP-Z (ZH), MAP-Z (UA), MAP-U (NW), MAP-U (CE) and MAP-U (SE). The figure shows for each item the standardized distance from the center of each cluster. This means that positive values occured more strongly than average, whereas negative values represented lower values compared to the mean behavior. **Notes.** MAP-Z (ZH): Mental Health Assessment of the Swiss Population in Zürich; MAP-Z (UA): Mental Health Assessment of the Ukrainian Population in Zürich; MAP-U (NW): Mental Health Assessment of the Population in Ukraine (North-West); MAP-U (CE): Mental Health Assessment of the Population in Ukraine (Central); MAP-U (SW): Mental Health Assessment of the Population in Ukraine (South-East); PHQ-15: Patient Health Questionaire-15; PHQ-9: GAD-7: Generalized Anxiety Disorder-7; Patient Health Questionaire-9; PCL-5: PTSD Checklist for DSM-5.

To further identify distinct symptom patterns across anxiety, depression, somatic and post-traumatic stress, cluster analysis was conducted. We conducted a k-means cluster analysis. For all five groups (MAP-Z (ZH), MAP-Z (UA) and MAP-U (SE)), 3 clusters were identified as optimal by the gap statistic. The clusters (**Figure 3**) demonstrated a clear differentiation in symptom severity across domains, as cluster 1 (high distress group), cluster 2 (moderate distress group) and cluster 3 (low distress group). We did not find symptom clusters according to the subgroups depression, anxiety, PTSD or somatic symptom cluster. Cluster 1 (high distress group) showed elevated scores across nearly all symptom domains, particularly for anxiety-related variables (e.g., afraid, worried), depressive symptoms (e.g., failure, tired), and PTSD stress reactivity (e.g., feel stress again, trouble falling asleep) and somatic complaints such as headache and fatigue. This identified pattern suggests a generalized distress cluster characterized by high comorbidity and symptom overlap across various symptom domains. Cluster 2 exhibited a more moderate symptom structure, with most symptom scores falling between the population mean and 0.5 SD above it. This group can be characterized as a moderate somatic-stress group not showing the global distress as seen in Cluster 1. Cluster 3 showed a low symptom burden across all domains, with all symptoms scoring consistently below the mean. This group can be therefore interpreted as a low-symptom cluster, reflecting low anxiety, depressive, somatic, and PTSD-related symptomatology.

**Figure 3.**
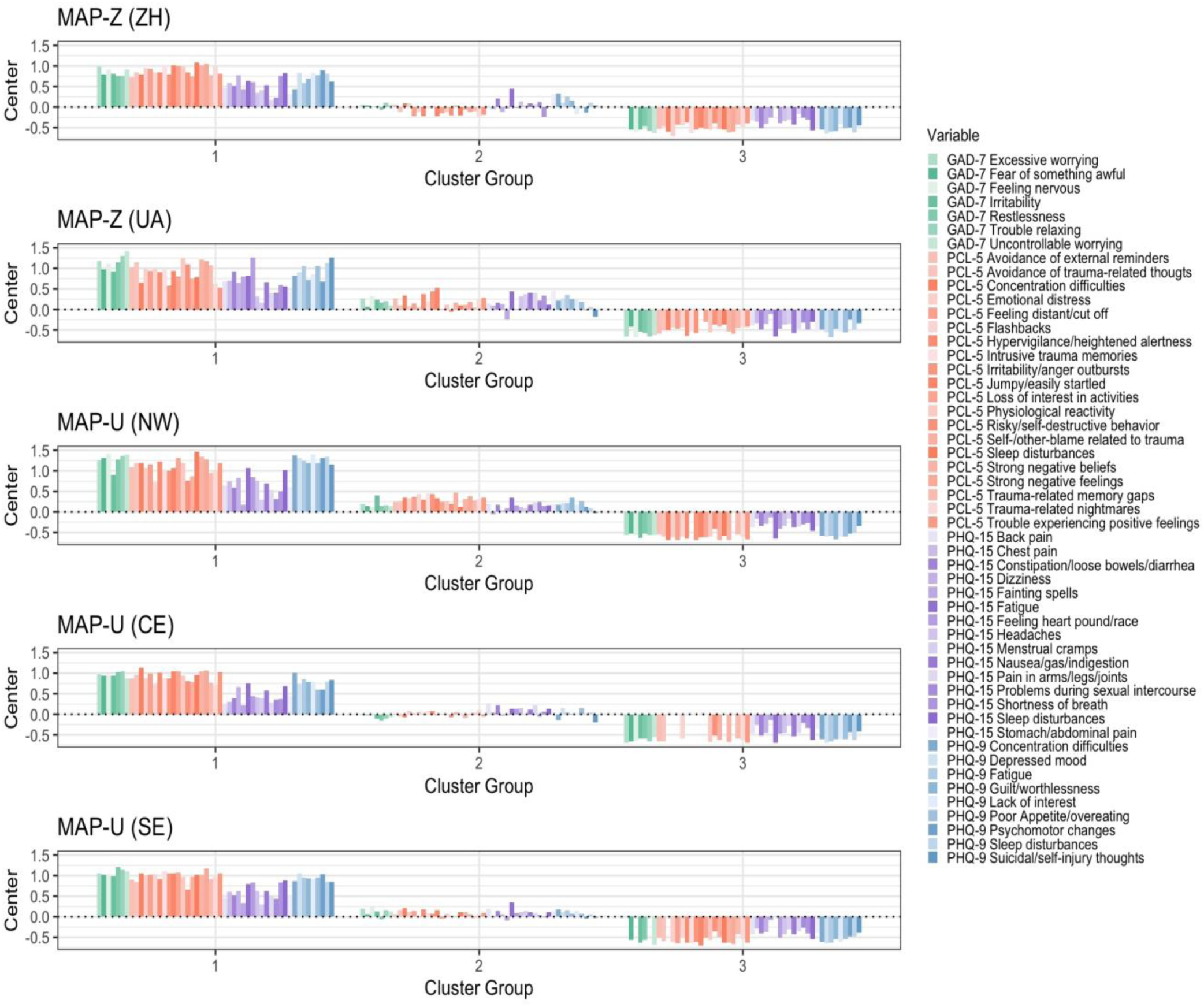
K-means cluster analysis (3 cluster) including PHQ-15, PHQ-9, GAD-7 and PCL-5 items for Swiss young adults living in Switzerland (MAP-Z (ZH)), young Ukrainian adults living in Switzerland (MAP-Z (UA)) and young Ukrainian adults in Ukraine (MAP-U (SE), MAP-U (CE), MAP-U (NW)). **Notes.** MAP-Z (ZH): Mental Health Assessment of the Swiss Population in Zürich; MAP-Z (UA): Mental Health Assessment of the Ukrainian Population in Zürich; MAP-U (NW): Mental Health Assessment of the Population in Ukraine (North-West); MAP-U (CE): Mental Health Assessment of the Population in Ukraine (Central); MAP-U (SW): Mental Health Assessment of the Population in Ukraine (South-East); PHQ-15: Patient Health Questionaire-15; PHQ-9: GAD-7: Generalized Anxiety Disorder-7; Patient Health Questionaire-9; PCL-5: PTSD Checklist for DSM-5.

We also applied the same cluster analysis approach to Ukrainian participants, both residing in Switzerland and living in different regions of Ukraine (also shown in **Figure 3**). This allowed us to compare patterns of symptom co-occurrence across populations with distinct stress exposures. Across all subsamples, a 3-cluster solution provided the best fit, consistently identifying a high-distress group (cluster 1), a moderate-distress group (cluster 2) and a low (more somatic) distress group (cluster 3). In addition, **Table 2**, shows the average symptom sum scores for the three mentioned clusters across all subgroups. For cluster 1, the average sum scores across all instruments and subgroups were high, above or close to the established screening cut-offs (as detailed in the methods section). For cluster 2, the average sum scores were lower than cluster 1 and higher than cluster 3 showing moderate symptom severities. In comparison, cluster 3 showed the lowest values with sum scores indicating mild symptom severity across all instruments.

**Table 2.** Average symptom sum scores for the three clusters across subgroups (MAP-Z (ZH), MAP-Z (UA), MAP-U (NW), MAP-U (CE) and MAP-U (SE)). **Notes.** MAP-Z (ZH): Mental Health Assessment of the Swiss Population in Zürich; MAP-Z (UA): Mental Health Assessment of the Ukrainian Population in Zürich; MAP-U (NW): Mental Health Assessment of the Population in Ukraine (North-West); MAP-U (CE): Mental Health Assessment of the Population in Ukraine (Central); MAP-U (SW): Mental Health Assessment of the Population in Ukraine (South-East); PHQ-15: Patient Health Questionaire-15; PHQ-9: GAD-7: Generalized Anxiety Disorder-7; Patient Health Questionaire-9; PCL-5: PTSD Checklist for DSM-5.

| Cluster | Instrument | MAP-Z<br>(ZH) | MAP-Z<br>(UA) | MAP-U<br>(NW) | MAP-U<br>(CE) | MAP-U<br>(SE) |
| --- | --- | --- | --- | --- | --- | --- |
| 1 | PHQ-15 | 12.1 | 15.9 | 15 | 13.9 | 15 |
| 1 | PHQ-9 | 13.4 | 18.8 | 20.1 | 16.4 | 18.2 |
| 1 | GAD-7 | 11.2 | 15.9 | 14.8 | 14 | 15.5 |
| 1 | PCL-5 | 33.2 | 51.6 | 50.7 | 49 | 51.8 |
| 2 | PHQ-15 | 8.2 | 12.2 | 10.2 | 10.6 | 10.4 |
| 2 | PHQ-9 | 8.8 | 12 | 10.4 | 9.7 | 10.8 |
| 2 | GAD-7 | 6.3 | 9.2 | 8 | 7.1 | 8.9 |
| 2 | PCL-5 | 13 | 32.9 | 31.7 | 26.1 | 29 |
| 3 | PHQ-15 | 3.7 | 5.5 | 5.5 | 5.5 | 5.3 |
| 3 | PHQ-9 | 4.1 | 6.3 | 3.8 | 4.4 | 5.2 |
| 3 | GAD-7 | 2.6 | 3.8 | 3.1 | 3.4 | 3.5 |
| 3 | PCL-5 | 4.4 | 15.5 | 9.1 | 9.1 | 12.4 |

**Notes.** MAP-Z (ZH): Mental Health Assessment of the Swiss Population in Zürich; MAP-Z (UA): Mental Health Assessment of the Ukrainian Population in Zürich; MAP-U (NW): Mental Health Assessment of the Population in Ukraine (North-West); MAP-U (CE): Mental Health Assessment of the Population in Ukraine (Central); MAP-U (SW): Mental Health Assessment of the Population in Ukraine (South-East); PHQ-15: Patient Health Questionnaire-15; PHQ-9: GAD-7: Generalized Anxiety Disorder-7; Patient Health Questionnaire-9; PCL-5: PTSD Checklist for DSM-5.

To assess the impact of the exclusion of participants with missing values in any of the items in the cluster analysis, we repeated the analysis two times where once all missing items were replaced by zero (the minimam possible value) and once by the maximum value possible for the respective item. Both cluster analyses resulted in a classification in a high-, moderate- and low-distress group that is comparable to our main results.

## Discussion

This study demonstrates that somatic and psychological symptoms in young adults cluster by overall symptom severity rather than by disorder-specific patterns. This pattern was remarkably consistent across populations with distinct cultural backgrounds and highly different stress exposures. Moreover, we observed moderate to strong correlations between somatic symptoms, depression, anxiety, and PTSD-related symptoms consistently across all subgroups of young adults. Rather than identifying discrete diagnostic clusters, we identified stable symptom severity-based patterns, forming three clusters (high distress, moderate distress and low distress cluster), across symptom dimensions for somatic, depression, anxiety and PTSD-related symptoms. Across all samples, higher somatic symptom burden was consistently associated with higher scores across all psychological symptom scales. These associations also emerged for non-overlapping diagnostic criteria, suggesting shared underlying mechanisms beyond diagnostic overlap. However, overlapping diagnostic items, such as items around sleep shared by the PHQ-15, PHQ-9 and PCL-5, items around fatigue shared by the PHQ-15 and PHQ-9, items around irritable behavior, nervous and anxious feelings shared by the GAD-7 and PCL-5, as well as trouble concentrating and loss of interest common to the PHQ-9 and PCL-5, may also partly contribute to the correlation, even more complicating differential diagnosis and treatment planning [4, 26, 27].

Previous research has documented the co-occurrence of somatic and psychological symptoms in both clinical and population-based samples [8, 21, 56–62]. Our study extends this literature by demonstrating this correlation at the population level for young adults persisting across diverse contexts. A strength of this study is the stability of the findings across three samples that differ substantially in both cultural context and, in particular, in levels of external stress exposure, including forced migration and war-related trauma experiences.

Although Swiss young adults and Ukrainian young adults living in Switzerland reported more preexisting mental health conditions than Ukrainians residing in Ukraine, descriptive analyses indicate a substantial burden of both somatic and psychological symptoms among Ukrainian young adults overall. Differences in reported chronic health conditions may reflect structural barriers, greater stigma, and limited access to mental health care in Ukraine, even prior to the escalation of the war, potentially leading to underdiagnosis of mental health conditions and delayed help-seeking [63, 64]. However, studies in Switzerland showed an overall high mental health burden among Swiss adolescents, particularly in relation to the impact of the COVID-19 pandemic, as well as the influence of social media use [16, 65–67]. Additionally, a vague or differing conceptualization of chronic mental health conditions may have influenced responses among Swiss and Ukrainian young adults [65]. The increased vulnerability of refugee populations to mental health disorders, linked to war exposure and pre-migration trauma as well as post-migration stressors (such as separation anxiety, resettlement challenges, and socioeconomic factors), may contribute to the higher prevalence of preexisting mental health conditions among Ukrainian refugees in Switzerland compared to Ukrainians residing in Ukraine [68–70].

Cluster analysis across all samples consistently revealed three distinct and replicable symptom severity patterns, ranging from low-distress symptom patterns to high-distress symptom patterns, reflecting generalized distress patterns with substantial symptom overlap and comorbidity. Importantly, no clusters aligned with specific diagnostic categories such as depression, anxiety, PTSD, or somatic symptom groups. Instead, symptom patterns reflected a continuum of symptom load multisystem distress, which is often insufficiently captured by traditional categorical diagnostic frameworks [26, 71].

These findings are consistent with transdiagnostic conceptualizations of psychopathology, including the HiTOP framework [34–36], reflecting shared underlying mechanisms. Multiple studies have associated the development and persistence of psychological and somatic symptoms with emotion regulation difficulties and stress reactivity [21, 45, 46, 66]. Somatic complaints are also prevalent in youth with emotional difficulties in general [67] and among those diagnosed with depression [68], trauma-related [69], and anxiety disorders [46]. Despite contextual variations, all subsamples demonstrated a continuum from low to high symptom burden overall three-cluster structure shows stability across the subsamples. The replication of this structure across culturally and contextually diverse populations further supports the robustness of these transdiagnostic processes and highlights the importance of considering somatic symptoms as integral components of youth mental health assessment and intervention. Given the cross-sectional nature of our baseline data, the directionality of this relation cannot be established at this stage. However, longitudinal research indicates bidirectional relationships, with somatic symptoms predicting anxiety and depression, supporting their role as early indicators of mental health challenges [10, 70–72].

Our findings also resonate with emerging diagnostic models such as Bodily Distress Disorder (BDD), which describe distressing bodily symptoms occurring alongside psychological burden [27, 73]. The observed symptom constellations also align with previous population-based estimates of Bodily Distress Syndrome (BDS) prevalence, ranging from 11.8% in Germany [74] to 16.1% in Denmark [75]. Moreover, multi-organ symptom presentations are associated with increased functional limitations, poorer self-rated health, and psychiatric comorbidity [75]. Current etiological concepts of somatic symptoms integrate both physical and mental processes and their complex interactions [21, 76]. Our results extend this evidence to young adulthood, a developmental period marked by heightened vulnerability and transition, and highlight the relevance of integrated, symptom-based classification models early in the life course.

The high overall and co-occurring symptom burden observed in young adults is concerning. Persistent somatic and psychological symptoms predict long-term psychiatric morbidity, reduced functioning, and increased healthcare utilization [10, 44]. Swiss cohort studies and long-term trends indicate heightened vulnerability to psychological distress and increasing symptom complaints among young adults [13, 14, 17, 42]. Together with our findings, this evidence highlights the importance of early identification, prevention, and intervention strategies that move beyond disorder-specific approaches and account for the complexity of multisymptomatic presentations in young adulthood. Taken together, these results underscore the urgent need for population-level prevention strategies and public health policies aimed at mitigating long-term mental health consequences in young adults.

### Strengths and Limitations

A key strength of this study is the inclusion of large and diverse samples of young adults based on population registries. The use of established and widely applied screening instruments (PHQ-9, GAD-7, PCL-5, PHQ-15) enhances comparability with prior research and clinical practice. Furthermore, the study extends previous population-based work by focusing specifically on young adults, a group underrepresented in somatic–psychological comorbidity research despite heightened vulnerability.

While this study provides valuable insights, there are several limitations. First, the cross-sectional analysis precludes causal inference regarding temporal relationships between somatic and psychological symptoms. Second, reliance on self-reported screening measures may introduce reporting bias and cannot replace clinical assessment; overlapping symptom content across scales may also inflate correlations (items around sleep shared by the PHQ-15, PHQ-9 and PCL-5, items around fatigue shared by the PHQ-15 and PHQ-9, items around irritable behavior, nervous and anxious feelings shared by the GAD-7 and PCL-5, as well as trouble concentrating and loss of interest common to the PHQ-9 and PCL-5). Third, differing recruitment strategies and contextual factors (e.g., displacement status, war exposure, potential digital divide) may influence symptom reporting and limit direct comparability across samples. Fourth, although cluster analysis supports latent subgroup identification, findings may vary with methodological choices, and replication using complementary analytical approaches (e.g., network analysis) is warranted.

The constraints related to the gender imbalance should be considered when interpreting how broadly the findings can be applied. However, the use of a random, population-based sampling strategy, along with substantial efforts to contact selected individuals, represents a notable strength.

## Conclusion

This study provides population-based evidence that somatic and psychological symptoms in young adults cluster by overall symptom severity rather than by diagnostic category, with a stable symptom structure observed across diverse stress-exposure contexts. These findings support dimensional, transdiagnostic approaches to mental health assessment and care in young adults and highlight the need for integrated, symptom-focused strategies to better capture the complexity of multisystem distress during this critical developmental period. Furthermore, our results underscore the importance of integrating somatic and psychological symptoms in prevention and early detection, by considering these domains jointly rather than as separate constructs.

## Data Availability

The datasets used and/or analyzed during the current study are available on reasonable request.

## List of abbreviations

BDD: Bodily Distress Disorder
BDS: Bodily Distress Syndrome
Cis: Confidence Intervals
DSM-5: Diagnostic and Statistical Manual of Mental Disorders, Fifth Edition
EBPI: Epidemioloy, Biostatistics and Prevention Institute
GAD-7: Generalized Anxiety Disorder-7
GBD: Global Burden of Disease
HiTOP: Hierarchical Taxonomy of Psychopathology
MAP: Mental Health Assessment of the Population
MAP-U: Mental Health Assessment of the Population in Ukraine
MAP-Z (UA): Mental Health Assessment of the Ukrainian Population in Zürich
MAP-Z (ZH): Mental Health Assessment of the Swiss Population in Zürich
MS: Multiple Sclerosis
ICD-11: International Classification of Diseases, 11^th^ Revision
NW: North-West
PCL-5: PTSD Checklist for DSM-5 PHQ-9: Patient Health Questionaire-9
PHQ-15: Patient Health Questionaire-15 PTSD: Posttraumatic Stress Disorder
REDCap: Research Electronic Data Capture
Scalex SP: Sample Size Calculator using Excel – Single Proportion
SD: Standard Deviation
SE: South-East
UA: Ukraine
UZH: University of Zürich
CE: Central

## Declarations

### Ethics approval and consent to participate

The study was approved by the responsible ethics committee of the canton of Zurich, Switzerland (BASEC-Nr. 2023-02247) as well as by the Commission on Bioethics of Sumy State University, Ukraine (ref-Nr. 60-0274), and electronic consent was obtained from all participants. The protocol has been prospectively registered on the International Standard Randomised Controlled Trial Number (ISRCTN17240415).

### Patient consent for publication

Consent obtained directly from patient(s).

### Patient and public involvement

Patients and/or the public were involved in the design, conduct, reporting or dissemination plans of this research. Refer to the Methods section for further details.

### Availability of data and materials

The datasets used and/or analysed during the current study are available on reasonable request.

### Study governance and partners

The project is led by the Epidemiology, Biostatistics and Prevention Institute (EBPI), University of Zurich (UZH), and Sumy State University, Ukraine.

### Authors’ contributions

VY, VvW, TB, SK, MK, AF, AK, AL, VP, SR and MAP contributed to the conceptualisation and methodology of the study. MK curated the data and provided support for the software. JB and AKS conducted the analysis of the data. VY, VP, VS, AL, AF, MB, OS and AK contributed to the investigation. VY, AF, and AK administered the project. VY, AF and AK contributed to the provision of resources. JB and AKS worked on the data visualisation. MK and AMB had full access to the raw data of the study. AKS and JB prepared the original draft. All authors contributed to reviewing and editing the manuscript. MAP handled the funding acquisition and supervision of the study. All authors have read and agreed to the published version of the manuscript.

## Acknowledgments

We thank the Swiss Federal Statistical Office for providing the list of randomly selected participants living in the canton of Zurich for the population-based part of our study. We also wish to thank all researchers and all staff that contributed to the implementation of MAP studies. We thank participants of the workshop “Mental Health Surveillance in Ukraine, an Essential Basis for Public Mental Health” which took place in March 2023 at the Collegium Helveticum (Zurich, Switzerland).

