## Supplementary material for "Distress above diagnostic constraints: transdiagnostic psychological and somatic symptom patterns in young adults": Figure S1, Table S1, Table S2-S6, Table S7, Figures S2-S4, Figures S7-S11

### Geographical overview of Ukrainian oblasts

To better reflect the different levels of war related stress exposure within Ukraine (MAP-U), we focused on three regional clusters formed from 10 selected oblasts across different parts of the country, each experiencing varying degrees of impact from the war (as described earlier [1, 2]). During the time of recruitment, the North-West cluster (NW) (Rivne, Lviv, and Chernivtsi oblasts) was located far from the war zone and faced relatively low security risks. The Central cluster (CE) (Zhytomyr, Kyiv, and Dnipropetrovsk oblasts) was closer to the border to Russia and experienced moderate security risks. The South-East cluster (SE) (Sumy, Kharkiv, Mykolaiv, and Kherson oblasts) was situated near the war zone and faced high security risks due to its close proximity to the Russian border. **Figure S1** maps a geographical overview about the 10 selected oblasts grouped in three clusters (NW, CE and SW) based on their geographical distance to the war border and level of direct involvement in ongoing war-activities.

**Figure S1.** Ukrainian oblasts included in the study and their clustering.

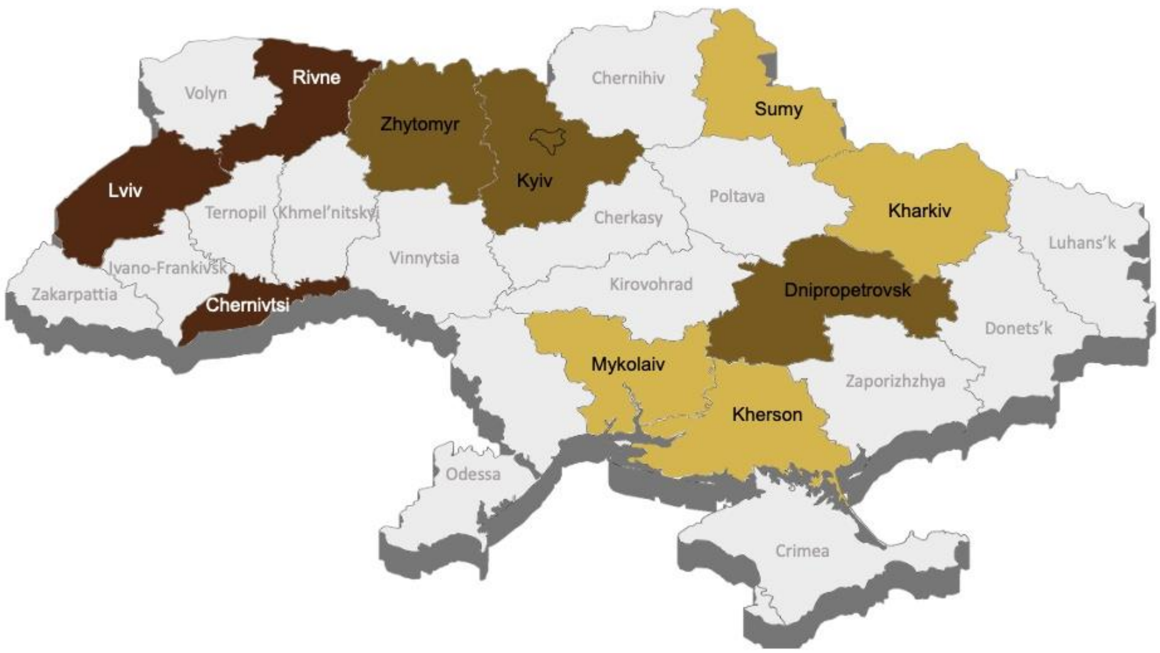

### Direct and indirect war-related experiences reported by Ukrainian sub-groups

In **Table S1** we analyzed the distinct experiences reported by Ukrainian participants from different regions, providing background information on their living conditions and assumed war-related stress exposures. We summarized war-related experiences reported that participants have been directly involved as well indirectly through their relatives.

**Table S1.** Direct and indirect war-related experiences reported by MAP-Z (UA) and MAP-U (NW, CE and SE) participants, given as number (percentage).

| | **MAP-Z (UA)** (n=91) | **MAP-U** (n=963) | | |
|---|---|---|---|---|
| | | North-West (n=211) | Central (n=230) | South-East (n=522) |
| Displacement experience[1] | 90 (98.9%)[2] | 70 (33.2%) | 85 (37.0%) | 291 (55.7%) |
| Residence damaged | 14 (15.4%) | 8 (3.8%) | 18 (7.8%) | 106 (20.3%) |
| Relatives mobilised | 12 (13.2%) | 63 (29.9%) | 73 (31.7%) | 120 (23.0%) |
| Injuries or trauma from hostilities[1] | 8 (8.8%) | 16 (7.6%) | 37 (16.1%) | 67 (12.8%) |
| Gender-based violence[1] | 3 (3.3%) | 6 (2.8%) | 6 (2.6%) | 17 (3.3%) |
| Death of relative | 14 (15.4%) | 34 (16.1%) | 24 (10.4%) | 67 (12.8%) |
| Separation from family members | 86 (94.5%) | 63 (29.9%) | 67 (29.1%) | 220 (42.1%) |

**Note.** [1]Direct and indirect experiences have been summarized. [2]Deviation from 100% may be explained by participants already living in Switzerland under different legal status or individual misunderstanding. Here presented as reported by participants for transparency. MAP-Z (UA): Mental Health Assessment of the Ukrainian Population in Zürich; MAP-U: Mental Health Assessment of the Population in Ukraine.

### Cross-tabulation of clinically relevant somatic symptoms (PHQ-15) and likely presence of depression, anxiety and post-traumatic stress disorder (PTDS)

We performed cross-tabulation analysis between psychological and somatic symptoms across all young adult subgroups. PHQ-15 scores range from 0 to 30, with an established cut-point of ≥10 reflecting moderate to severe somatic symptom severity [3]. This cut-off was identified as optimal to predict the diagnosis of a somatoform disorder in the primary care setting, resulting in a sensitivity of 0.80 and specificity of 0.59 [4, 5]. The moderate specificity reflects its design as a broad screening instrument for common somatic symptoms rather than a diagnostic tool. By capturing a wide range of physical complaints that frequently occur in outpatient and community settings, the PHQ-15 prioritizes sensitivity, which is appropriate for population-based research [4, 5]. PHQ-9 scores range from 0–27, with a cut-point of ≥10 indicating moderate-to-severe depression (sensitivity 0.88, specificity 0.88) [6, 7]. For anxiety, a GAD-7 cut-point of ≥10 was applied (sensitivity 0.89, specificity 0.82) [8]. PTSD symptoms were dichotomized using a PCL-5 cut-point of ≥31 (sensitivity 0.94, specificity 0.94) [9].

The results indicate a high comorbidity of clinically relevant (defined as cut-off ≥10) somatic symptoms with depression, anxiety, and PTSD among all study arms (MAP-Z (ZH), MAP-Z (UA), MAP-U (NW), MAP-U (CE) and MAP-U (SE)) (**Tables S2-S6**).

**Table S2.** Cross-tabulation of clinically relevant somatic symptoms and likely presence of depression, anxiety and PTSD among MAP-Z (ZH).

| | **PTSD (PCL-5)** | |
|---|---|---|
| | **0**<br>N = 108[1] | **1**<br>N = 19[1] |
| Anxiety (GAD-7) | | |
| 0 | 92 (85%) | 6 (32%) |
| 1 | 16 (15%) | 13 (68%) |

| | | |
|---|---|---|
| Depression (PHQ-9) | | |
| 0 | 76 (70%) | 3 (16%) |
| 1 | 32 (30%) | 16 (84%) |
| [1] n (%) | | |

| | **Anxiety (GAD-7)** | |
|---|---|---|
| | **0**<br>N = 98[1] | **1**<br>N = 29[1] |
| Depression (PHQ-9) | | |
| 0 | 72 (73%) | 7 (24%) |
| 1 | 26 (27%) | 22 (76%) |
| [1] n (%) | | |

| | **Somatic Symptoms (PHQ-15)** | |
|---|---|---|
| | **0**<br>N = 90[1] | **1**<br>N = 37[1] |
| PTSD (PCL-5) | | |
| 0 | 81 (90%) | 27 (73%) |
| 1 | 9 (10%) | 10 (27%) |
| Anxiety (GAD-7) | | |
| 0 | 80 (89%) | 18 (49%) |
| 1 | 10 (11%) | 19 (51%) |
| Depression (PHQ-9) | | |
| 0 | 65 (72%) | 14 (38%) |
| 1 | 25 (28%) | 23 (62%) |
| [1] n (%) | | |

**Table S3.** Cross-tabulation of clinically relevant somatic symptoms and likely presence of depression, anxiety and PTSD among MAP-Z (UA).

| | **PTSD (PCL-5)** | |
|---|---|---|
| | **0**<br>N = 56[1] | **1**<br>N = 35[1] |
| Anxiety (GAD-7) | | |

| | | |
|---|---|---|
| 0 | 50 (89%) | 14 (40%) |
| 1 | 6 (11%) | 21 (60%) |
| Depression (PHQ-9) | | |
| 0 | 38 (68%) | 3 (8.6%) |
| 1 | 18 (32%) | 32 (91%) |
| [1] n (%) | | |

| | **Anxiety (GAD-7)** | |
|---|---|---|
| | **0**<br>N = 64[1] | **1**<br>N = 27[1] |
| Depression (PHQ-9) | | |
| 0 | 37 (58%) | 4 (15%) |
| 1 | 27 (42%) | 23 (85%) |
| [1] n (%) | | |

| | **Somatic Symptoms (PHQ-15)** | |
|---|---|---|
| | **0**<br>N = 45[1] | **1**<br>N = 46[1] |
| PTSD (PCL-5) | | |
| 0 | 39 (87%) | 17 (37%) |
| 1 | 6 (13%) | 29 (63%) |
| Anxiety (GAD-7) | | |
| 0 | 40 (89%) | 24 (52%) |
| 1 | 5 (11%) | 22 (48%) |
| Depression (PHQ-9) | | |
| 0 | 33 (73%) | 8 (17%) |
| 1 | 12 (27%) | 38 (83%) |
| [1] n (%) | | |

**Table S4.** Cross-tabulation of clinically relevant somatic symptoms and likely presence of depression, anxiety and PTSD among MAP-U (NW).

| | **PTSD (PCL-5)** | |
|---|---|---|
| | **0** | **1** |

| | N = 136[1] | N = 75[1] |
|---|---|---|
| Anxiety (GAD-7) | | |
| 0 | 126 (93%) | 33 (44%) |
| 1 | 10 (7.4%) | 42 (56%) |
| Depression (PHQ-9) | | |
| 0 | 112 (82%) | 15 (20%) |
| 1 | 24 (18%) | 60 (80%) |
| [1] n (%) | | |

| | **Anxiety (GAD-7)** | |
|---|---|---|
| | **0**<br>N = 159[1] | **1**<br>N = 52[1] |
| Depression (PHQ-9) | | |
| 0 | 118 (74%) | 9 (17%) |
| 1 | 41 (26%) | 43 (83%) |
| [1] n (%) | | |

| | **Somatic Symptoms (PHQ-15)** | |
|---|---|---|
| | **0**<br>N = 128[1] | **1**<br>N = 83[1] |
| PTSD (PCL-5) | | |
| 0 | 101 (79%) | 35 (42%) |
| 1 | 27 (21%) | 48 (58%) |
| Anxiety (GAD-7) | | |
| 0 | 110 (86%) | 49 (59%) |
| 1 | 18 (14%) | 34 (41%) |
| Depression (PHQ-9) | | |
| 0 | 101 (79%) | 26 (31%) |
| 1 | 27 (21%) | 57 (69%) |
| [1] n (%) | | |

**Table S5.** Cross-tabulation of clinically relevant somatic symptoms and likely presence of depression, anxiety and PTSD among MAP-U (CE).

| | **PTSD (PCL-5)** | |
|---|---|---|
| | **0** | **1** |
| | N = 145[1] | N = 85[1] |
| Anxiety (GAD-7) | | |
| 0 | 125 (86%) | 26 (31%) |
| 1 | 20 (14%) | 59 (69%) |
| Depression (PHQ-9) | | |
| 0 | 112 (77%) | 15 (18%) |
| 1 | 33 (23%) | 70 (82%) |
| [1] n (%) | | |

| | **Anxiety (GAD-7)** | |
|---|---|---|
| | **0** | **1** |
| | N = 151[1] | N = 79[1] |
| Depression (PHQ-9) | | |
| 0 | 112 (74%) | 15 (19%) |
| 1 | 39 (26%) | 64 (81%) |
| [1] n (%) | | |

| | **Somatic Symptoms (PHQ-15)** | |
|---|---|---|
| | **0** | **1** |
| | N = 116[1] | N = 114[1] |
| PTSD (PCL-5) | | |
| 0 | 98 (84%) | 47 (41%) |
| 1 | 18 (16%) | 67 (59%) |
| Anxiety (GAD-7) | | |
| 0 | 100 (86%) | 51 (45%) |
| 1 | 16 (14%) | 63 (55%) |
| Depression (PHQ-9) | | |
| 0 | 92 (79%) | 35 (31%) |
| 1 | 24 (21%) | 79 (69%) |
| [1] n (%) | | |

**Table S6.** Cross-tabulation of clinically relevant somatic symptoms and likely presence of depression, anxiety and PTSD among MAP-U (SE).

| | **PTSD (PCL-5)** | |
|---|---|---|
| | **0**<br>N = 327[1] | **1**<br>N = 195[1] |
| Anxiety (GAD-7) | | |
| 0 | 282 (86%) | 57 (29%) |
| 1 | 45 (14%) | 138 (71%) |
| Depression (PHQ-9) | | |
| 0 | 242 (74%) | 37 (19%) |
| 1 | 85 (26%) | 158 (81%) |
| [1] n (%) | | |

| | **Anxiety (GAD-7)** | |
|---|---|---|
| | **0**<br>N = 339[1] | **1**<br>N = 183[1] |
| Depression (PHQ-9) | | |
| 0 | 249 (73%) | 30 (16%) |
| 1 | 90 (27%) | 153 (84%) |
| [1] n (%) | | |

| | **Somatic Symptoms (PHQ-15)** | |
|---|---|---|
| | **0**<br>N = 288[1] | **1**<br>N = 234[1] |
| PTSD (PCL-5) | | |
| 0 | 236 (82%) | 91 (39%) |
| 1 | 52 (18%) | 143 (61%) |
| Unknown | 3 | 1 |
| Anxiety (GAD-7) | | |
| 0 | 236 (82%) | 103 (44%) |
| 1 | 52 (18%) | 131 (56%) |
| Depression (PHQ-9) | | |
| 0 | 213 (74%) | 66 (28%) |
| 1 | 75 (26%) | 168 (72%) |

[1] n (%)

### Frequency of the items of the PHQ-15

**Figures S2-S4** show the frequency of the single items of the PHQ-15 items reported in the Zurich (MAP-Z (ZH), MAP-Z (UA)) and Ukrainian (MAP-U (NW), MAP-U (CE) and MAP-U (SE)) samples. The most frequently reported somatic complaints among all sub-groups were fatigue, sleep difficulties, back pain and headache. For the Ukrainian sub-groups in Switzerland as well as in Ukraine (**Figures S3-S6**) the symptoms heart racing and shortness of breath was more frequent reported. A stacked bar plot of PHQ-15 item responses revealed considerable heterogeneity in symptom expression.

**Figure S2**. Frequency of the single items of the PHQ-15 items in MAP-Z (ZH).

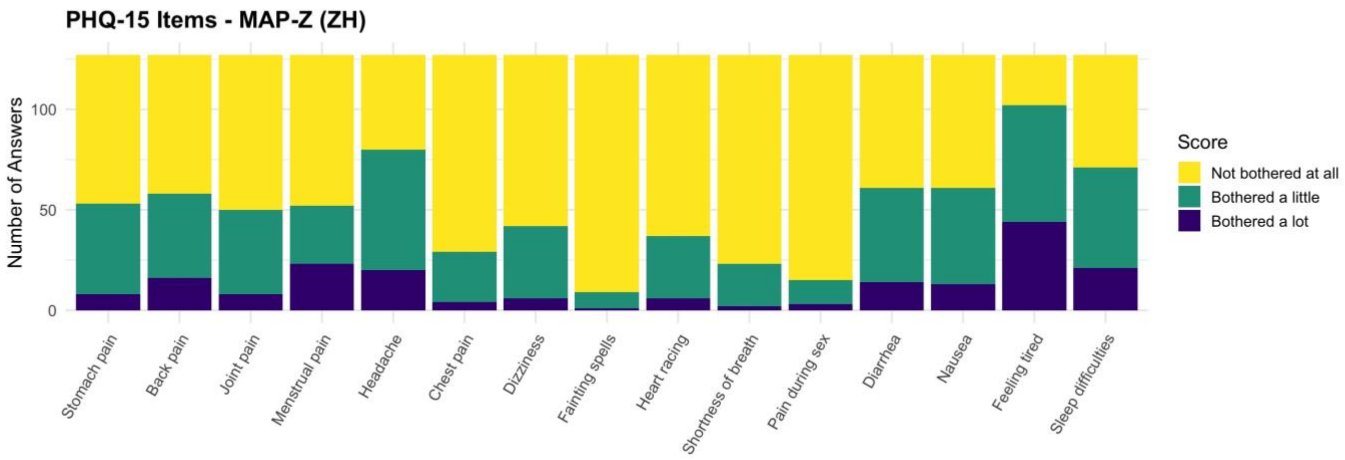

**Figure S3**. Frequency of the single items of the PHQ-15 items in MAP-Z (UA).

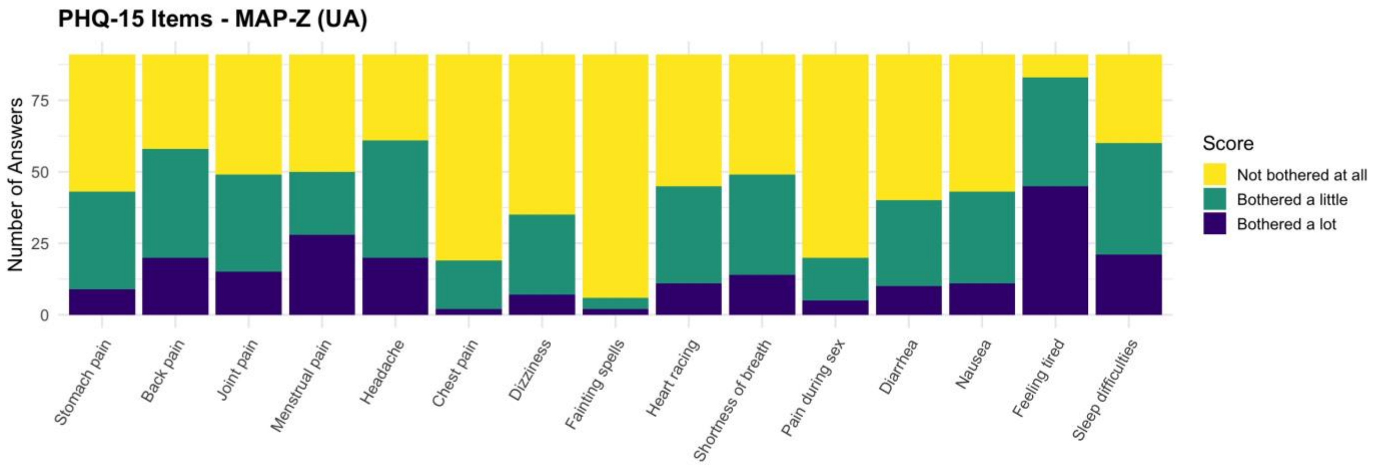

**Figure S4**. Frequency of the single items of the PHQ-15 items in MAP-U (NW).

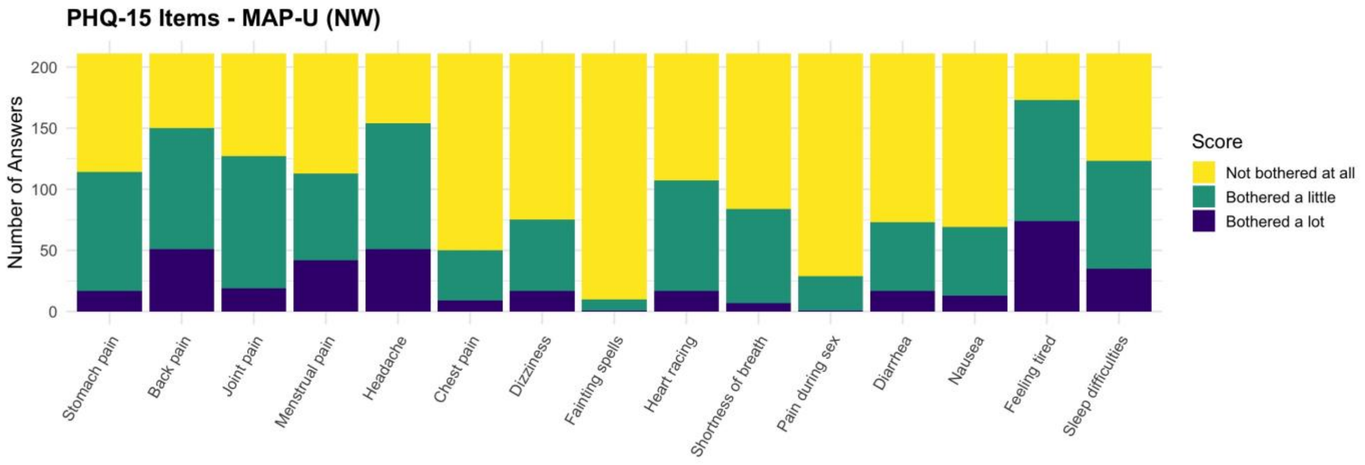

**Figure S5**. Frequency of the single items of the PHQ-15 items in MAP-U (CE).

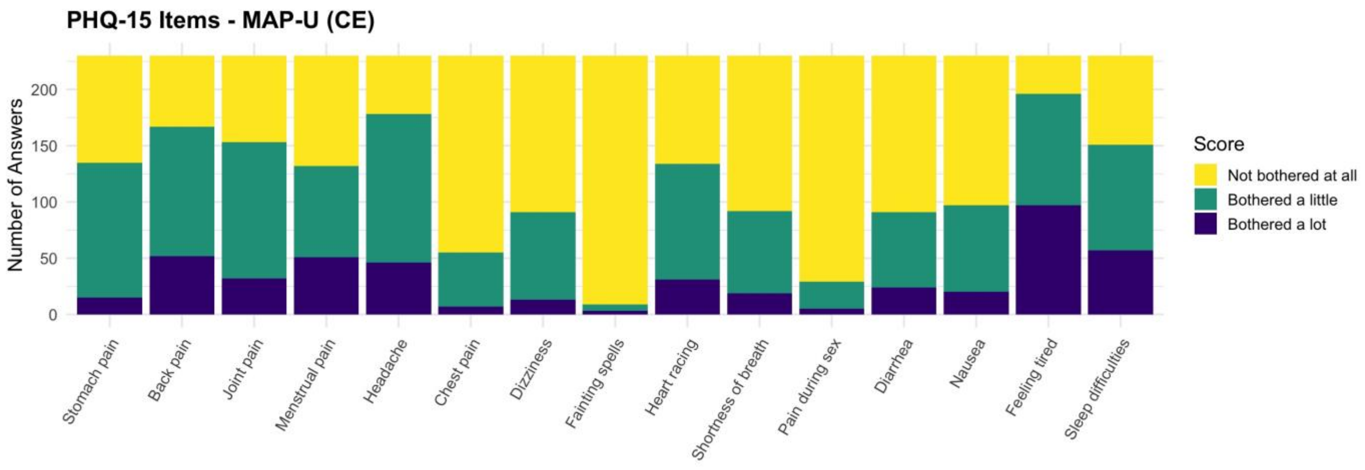

**Figure S6**. Frequency of the single items of the PHQ-15 items in MAP-U (SE).

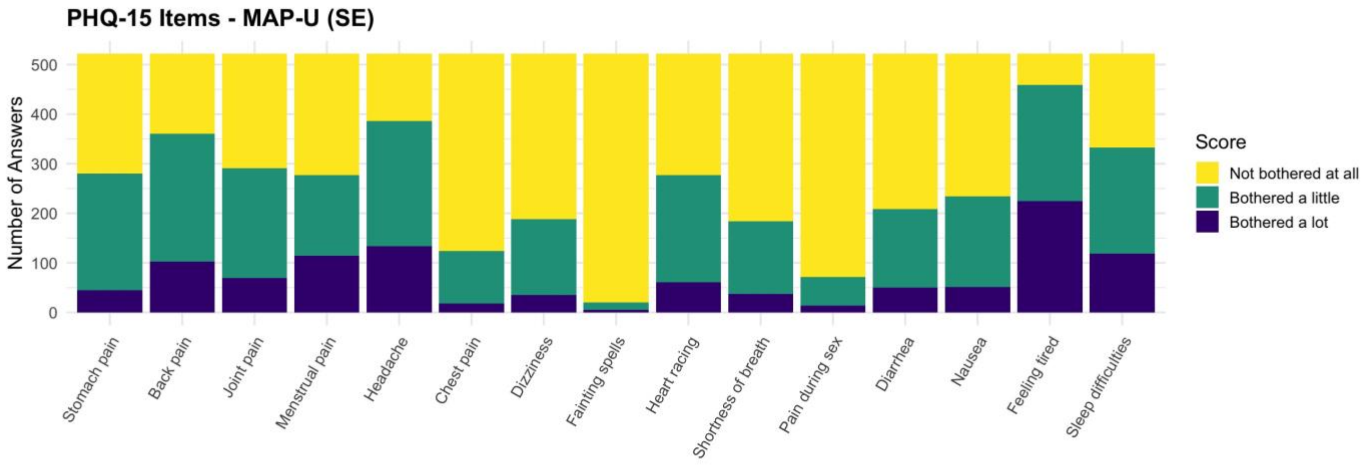

### Symptom burden (PHQ-15, PHQ-9, GAD-7 and PCL-5 sum scores) by study cohorts and oblasts (MAP-U)

**Table S7** illustrates the corresponding descriptive statistics. For each group, the table reports the median, the first and third quartiles (Q1 and Q3), and the minimum and maximum non-outlier values.

**Table S7**. PHQ-15, PHQ-9, GAD-7 and PCL-5 sum scores by study cohorts.

| Outcome | Study arm | n | min. | Q1 | median | Q3 | max. |
|---|---|---|---|---|---|---|---|
| PHQ-15 | MAP-Z (ZH) | 127 | 0 | 4 | 6 | 10 | 22 |

| | | | | | | | |
|---|---|---|---|---|---|---|---|
| PHQ-15 | MAP-Z (UA) | 91 | 2 | 5 | 10 | 13 | 30 |
| PHQ-15 | MAP-U (NW) | 211 | 0 | 5 | 8 | 12 | 23 |
| PHQ-15 | MAP-U (CE) | 230 | 0 | 6 | 9 | 13 | 22 |
| PHQ-15 | MAP-U (SE) | 522 | 0 | 5 | 9 | 13 | 27 |
| PHQ-9 | MAP-Z (ZH) | 127 | 0 | 4 | 8 | 11 | 22 |
| PHQ-9 | MAP-Z (UA) | 91 | 0 | 6 | 10 | 14 | 25 |
| PHQ-9 | MAP-U (NW) | 211 | 0 | 4 | 8 | 12 | 27 |
| PHQ-9 | MAP-U (CE) | 230 | 0 | 5 | 8 | 13.8 | 27 |
| PHQ-9 | MAP-U (SE) | 522 | 0 | 5 | 9 | 14 | 27 |
| GAD-7 | MAP-Z (ZH) | 127 | 0 | 2.5 | 5 | 9 | 19 |
| GAD-7 | MAP-Z (UA) | 91 | 0 | 4 | 7 | 11.5 | 21 |
| GAD-7 | MAP-U (NW) | 211 | 0 | 3 | 6 | 9 | 21 |
| GAD-7 | MAP-U (CE) | 230 | 0 | 4 | 6 | 11 | 21 |
| GAD-7 | MAP-U (SE) | 522 | 0 | 4 | 7 | 12 | 21 |
| PCL-5 | MAP-Z (ZH) | 127 | 0 | 4 | 11 | 24 | 65 |
| PCL-5 | MAP-Z (UA) | 91 | 0 | 15.5 | 26 | 39 | 60 |
| PCL-5 | MAP-U (NW) | 211 | 0 | 10 | 22 | 36 | 67 |
| PCL-5 | MAP-U (CE) | 230 | 0 | 11 | 23 | 37 | 80 |
| PCL-5 | MAP-U (SE) | 522 | 0 | 14 | 24 | 38.8 | 80 |

### Descriptive results of PHQ-15 items stratified for symptom burden of PHQ-9, GAD-7 and PCL-5

**Figures S7-S11** present the distribution of depression (PHQ-9), anxiety (GAD-7), and PTSD (PCL-5) scores by severity of each somatic symptom (PHQ-15 items) across the three PHQ-15 response categories («not bothered at all», «bothered a little», «bothered a lot») for each of the 15 somatic symptoms among all sub-groups. In all sub-groups across all three mental health outcomes, higher somatic symptom severity was associated with higher psychological symptom scores. Participants reported being «bothered a lot» by a given somatic symptom generally exhibited the highest median scores for depression, anxiety, and PTSD. Overall, these analyses reveal a consistent, symptom-specific graded association between the severity of somatic complaints and the burden of psychological symptoms.

**Figure S7.** Boxplots of PHQ-9, GAD-7 and PCL-5, stratified by somatic symptom severity for MAP-Z (ZH).

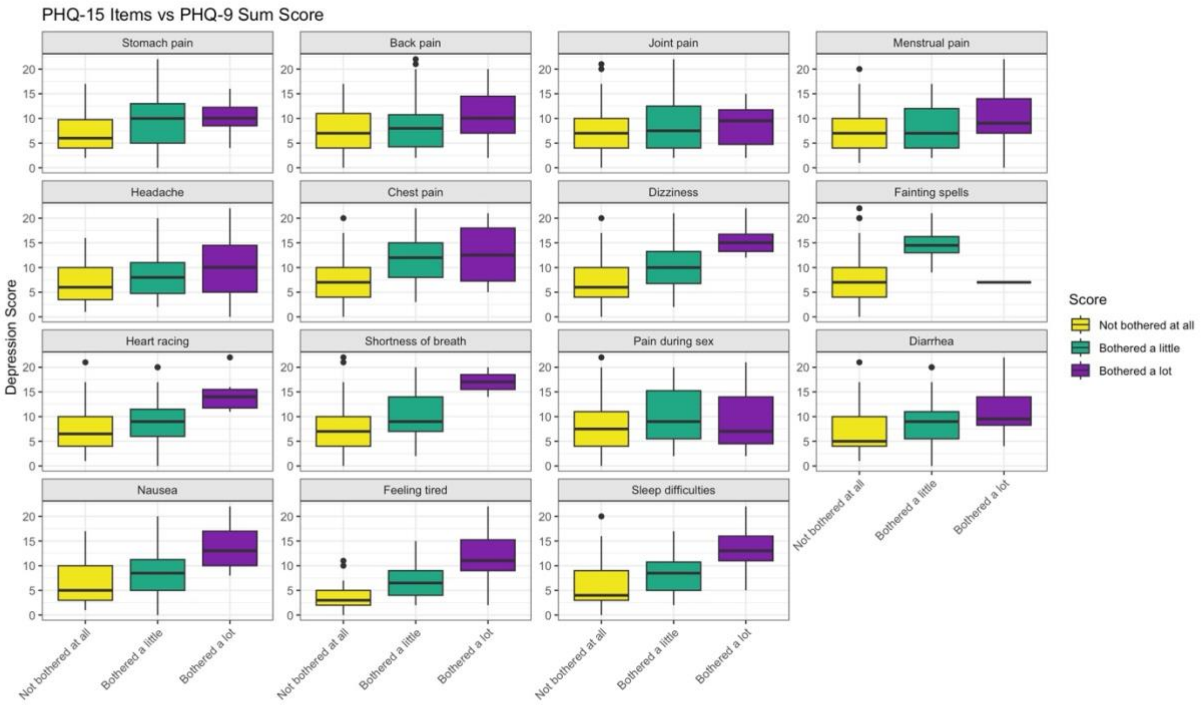

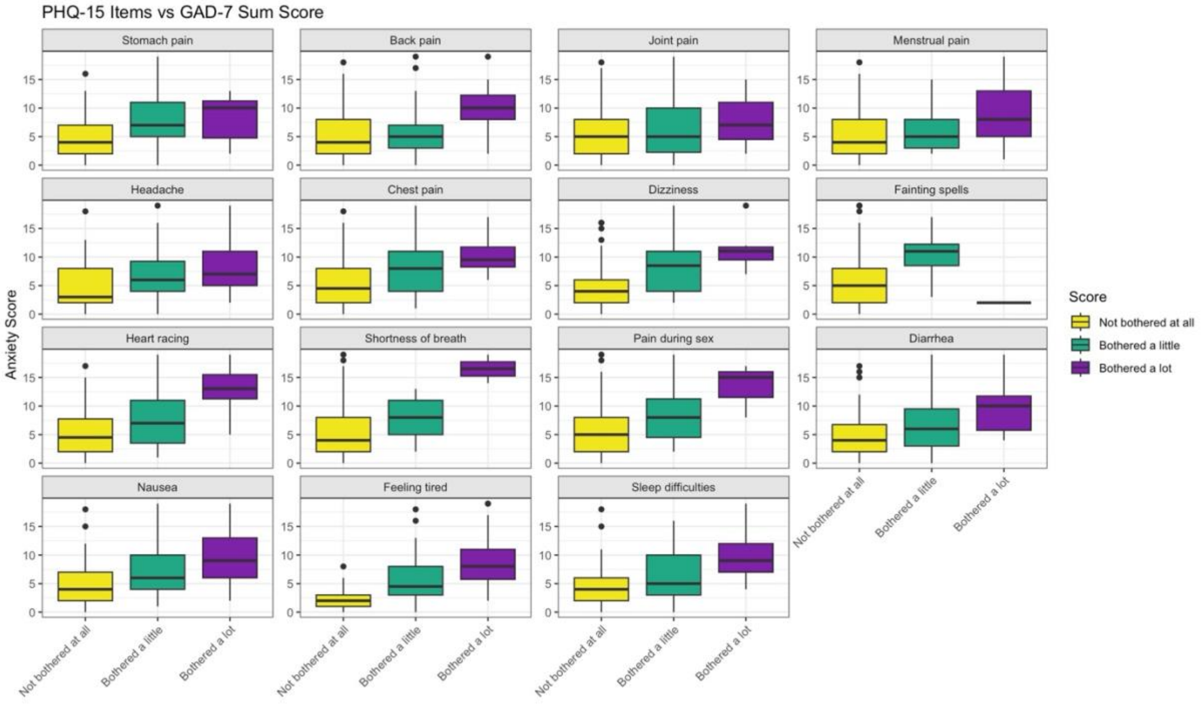

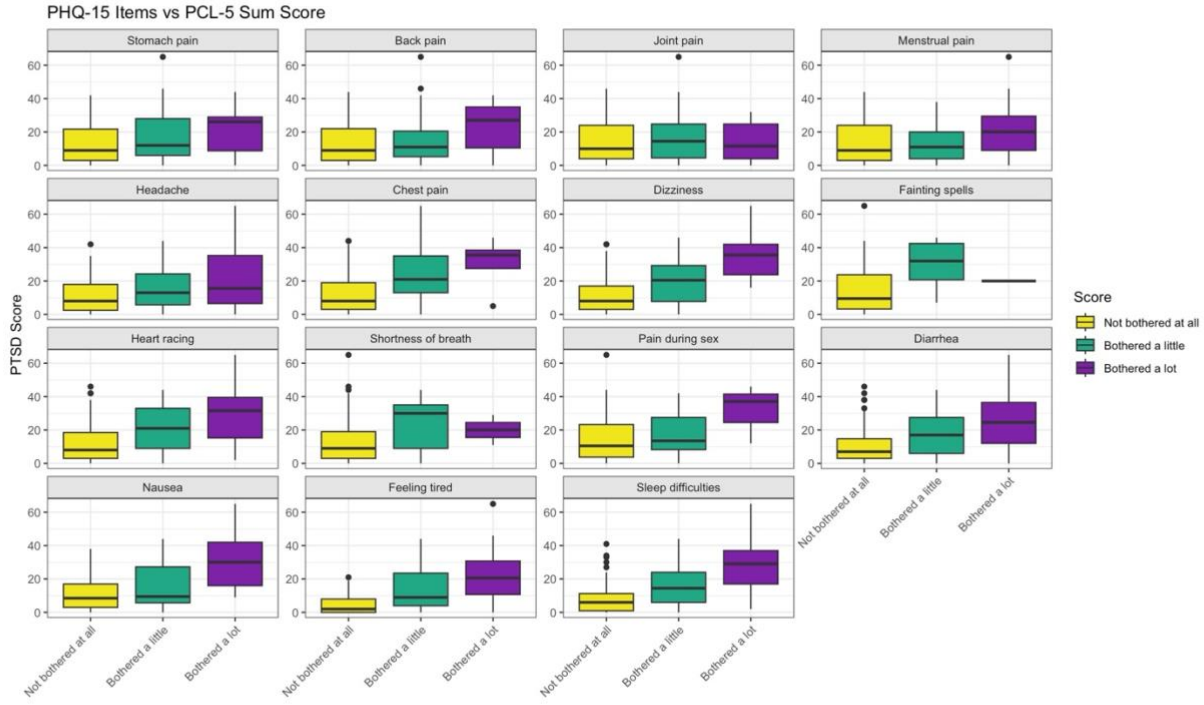

**Figure S8**. Boxplots of PHQ-9, GAD-7 and PCL-5, stratified by somatic symptom severity for MAP-Z (UA).

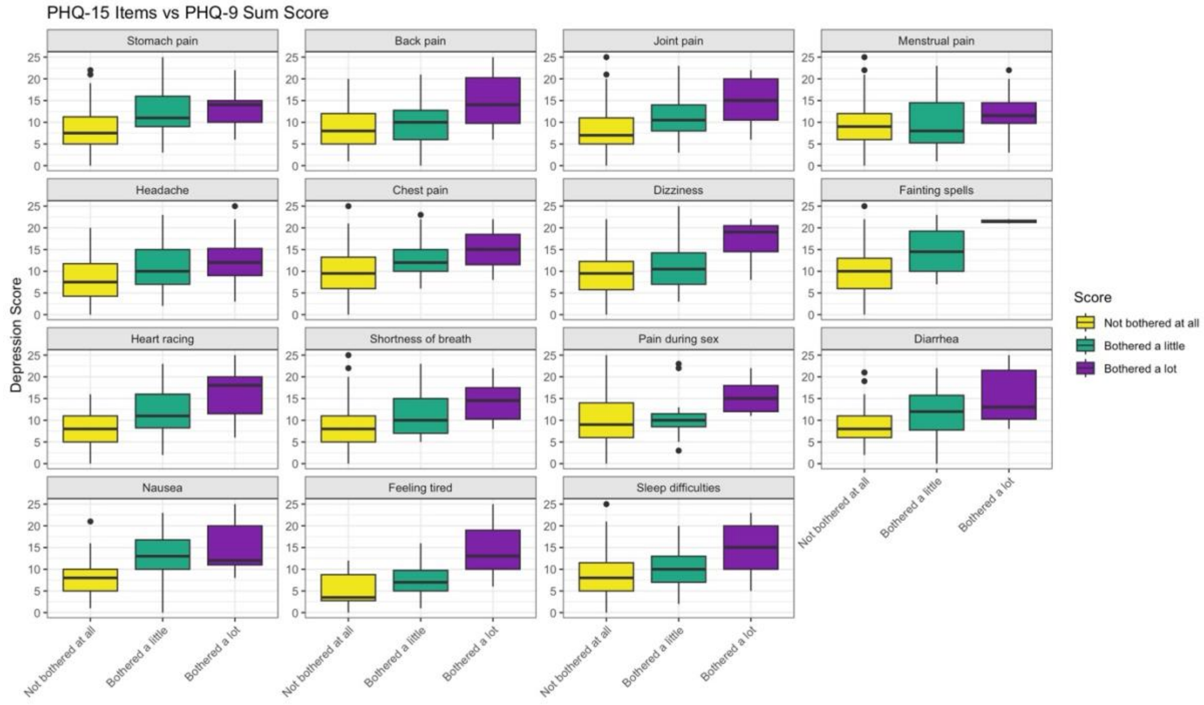

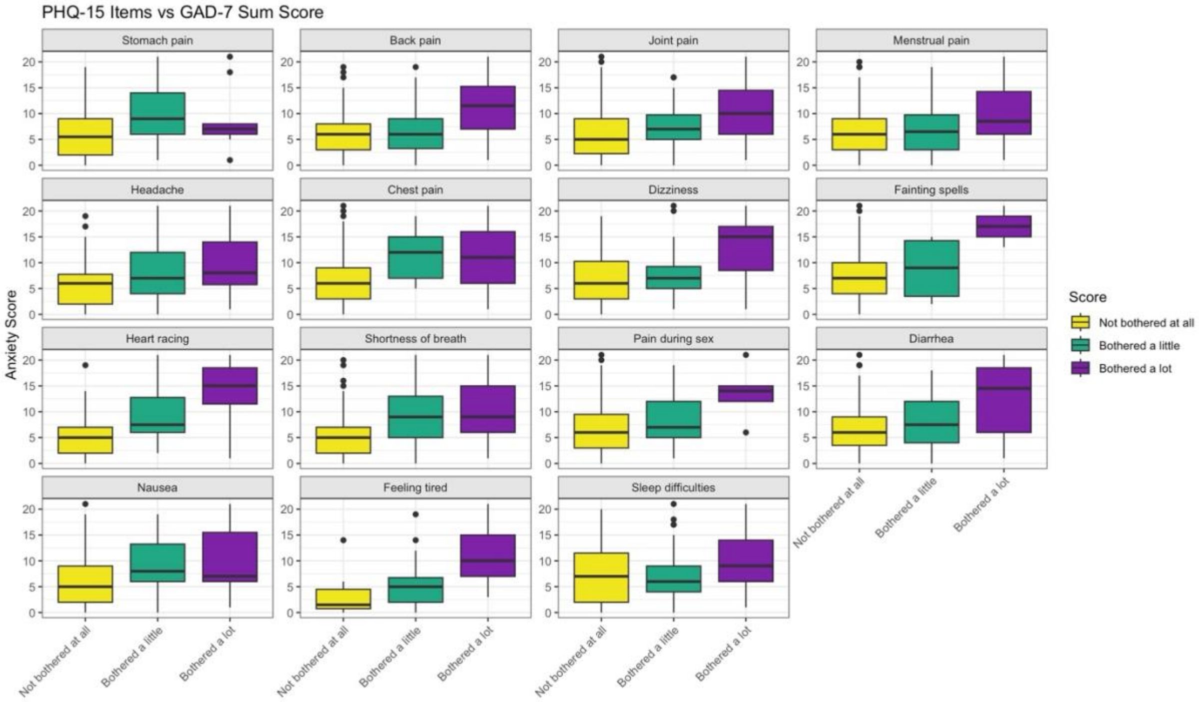

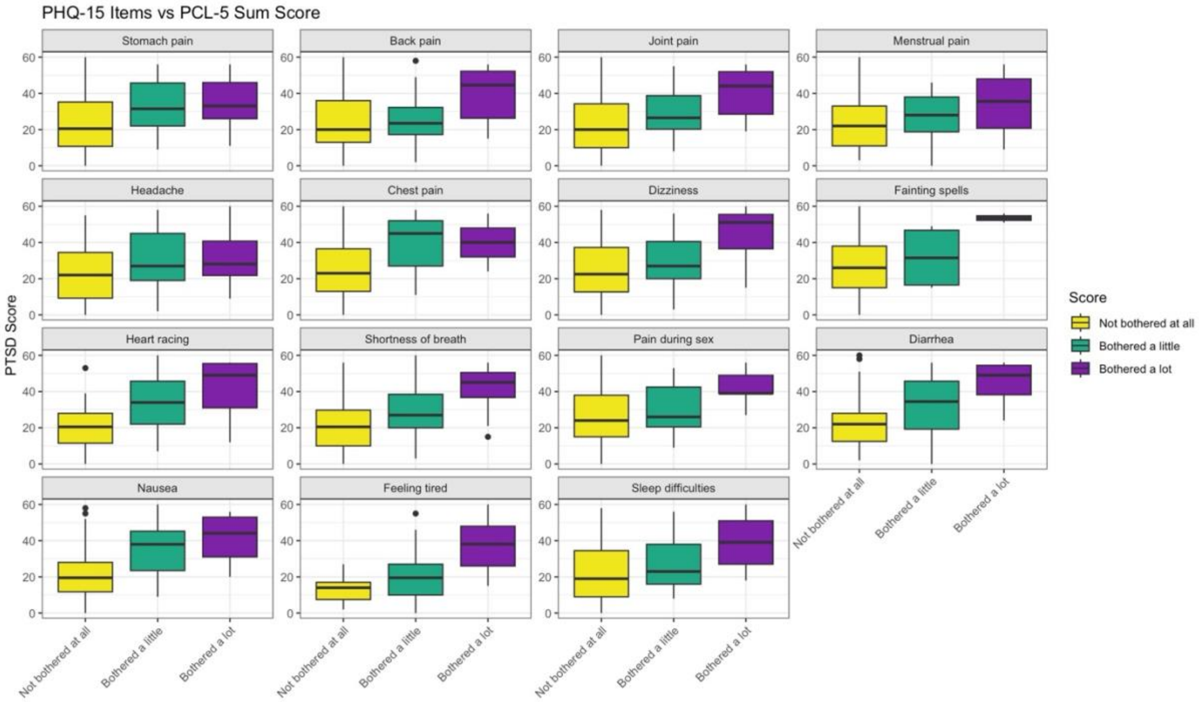

**Figure S9**. Boxplots of PHQ-9, GAD-7 and PCL-5, stratified by somatic symptom severity for MAP-U (NW).

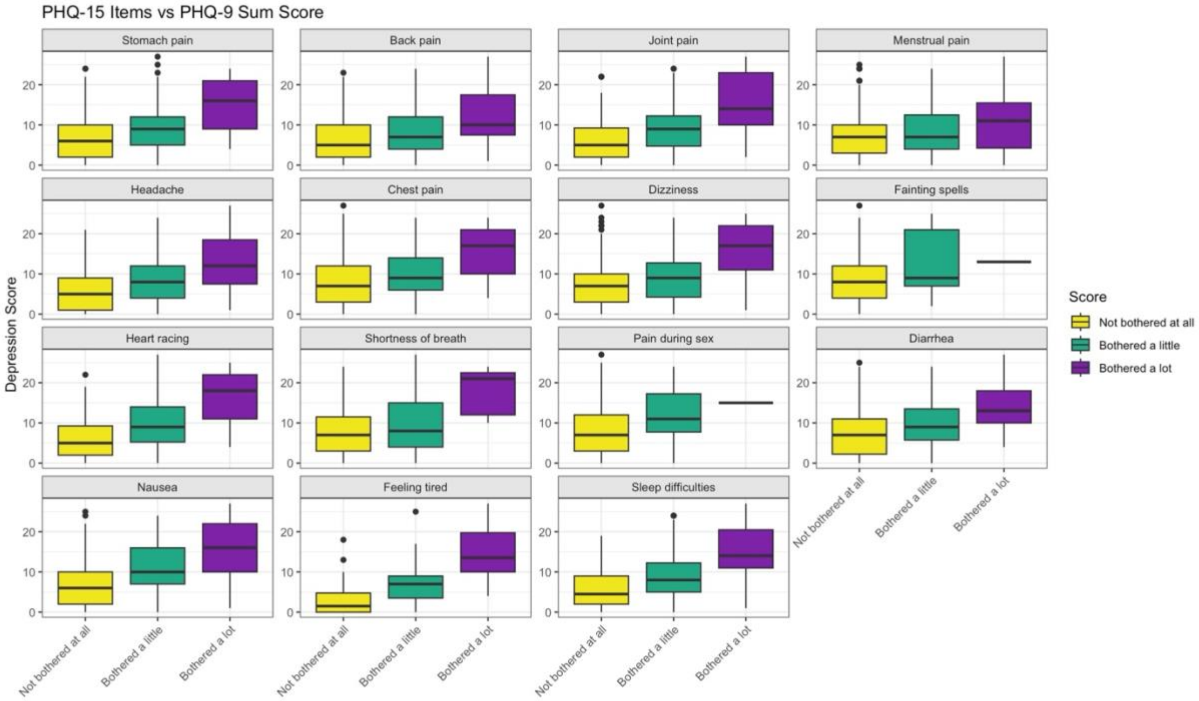
PHQ-15 Items vs PHQ-9 Sum Score
Stomach pain
Back pain
Joint pain
Menstrual pain
Headache
Chest pain
Dizziness
Fainting spells
Heart racing
Shortness of breath
Pain during sex
Diarrhea
Nausea
Feeling tired
Sleep difficulties
Depression Score
Score
Not bothered at all
Bothered a little
Bothered a lot

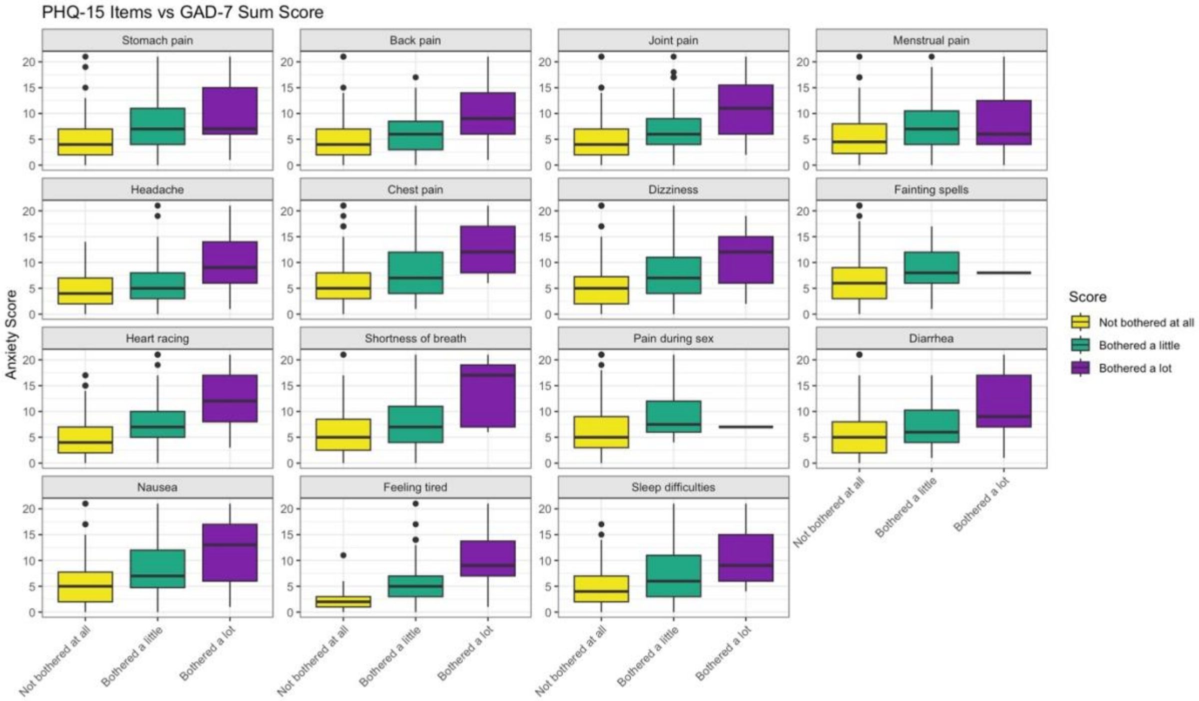
PHQ-15 Items vs GAD-7 Sum Score
Stomach pain
Back pain
Joint pain
Menstrual pain
Headache
Chest pain
Dizziness
Fainting spells
Heart racing
Shortness of breath
Pain during sex
Diarrhea
Nausea
Feeling tired
Sleep difficulties
Anxiety Score
Score
Not bothered at all
Bothered a little
Bothered a lot

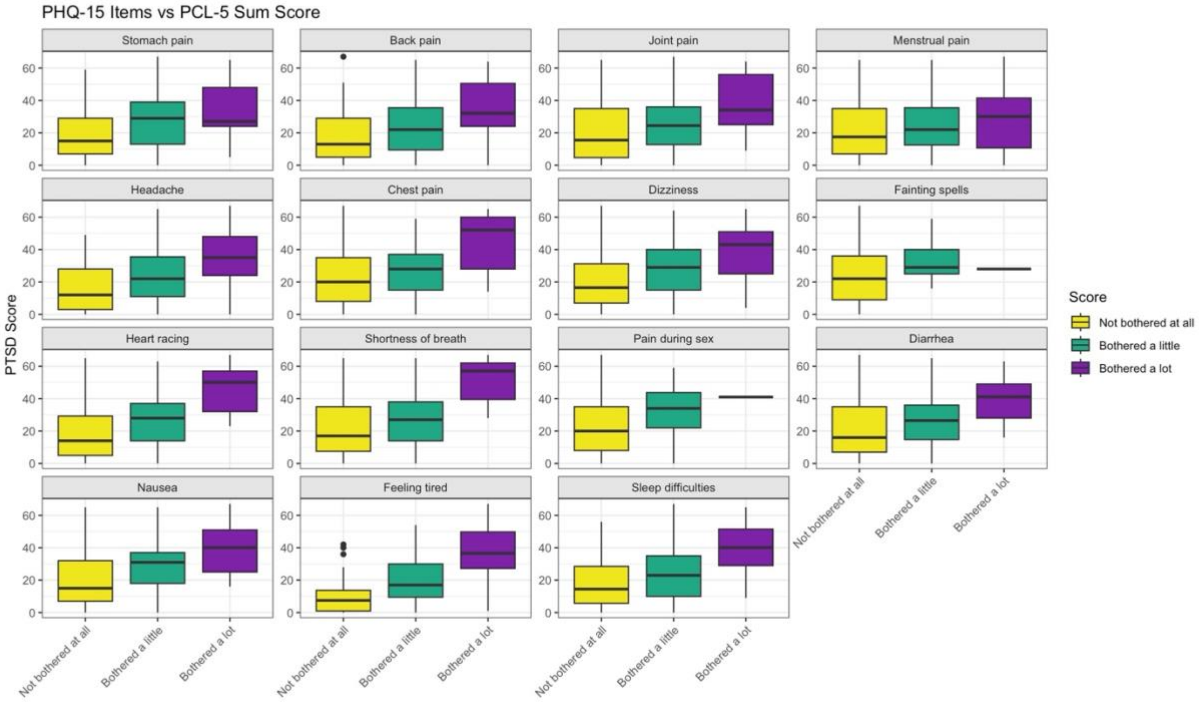

**Figure S10**. Boxplots of PHQ-9, GAD-7 and PCL-5, stratified by somatic symptom severity for MAP-U (CE).

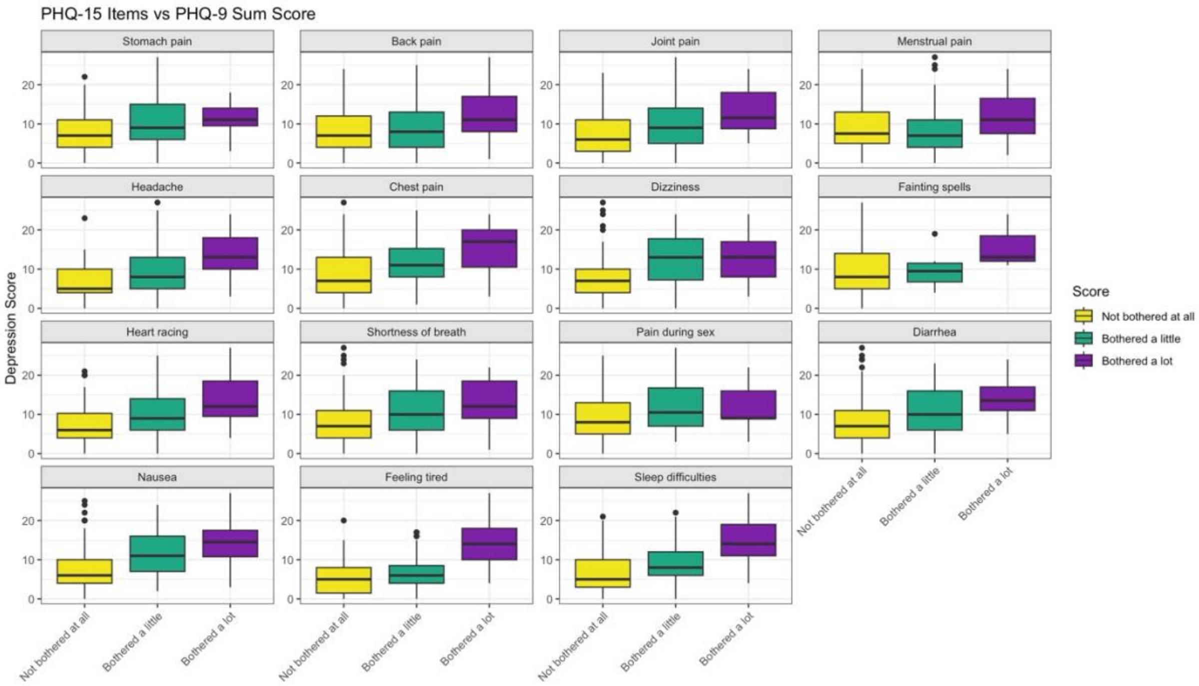

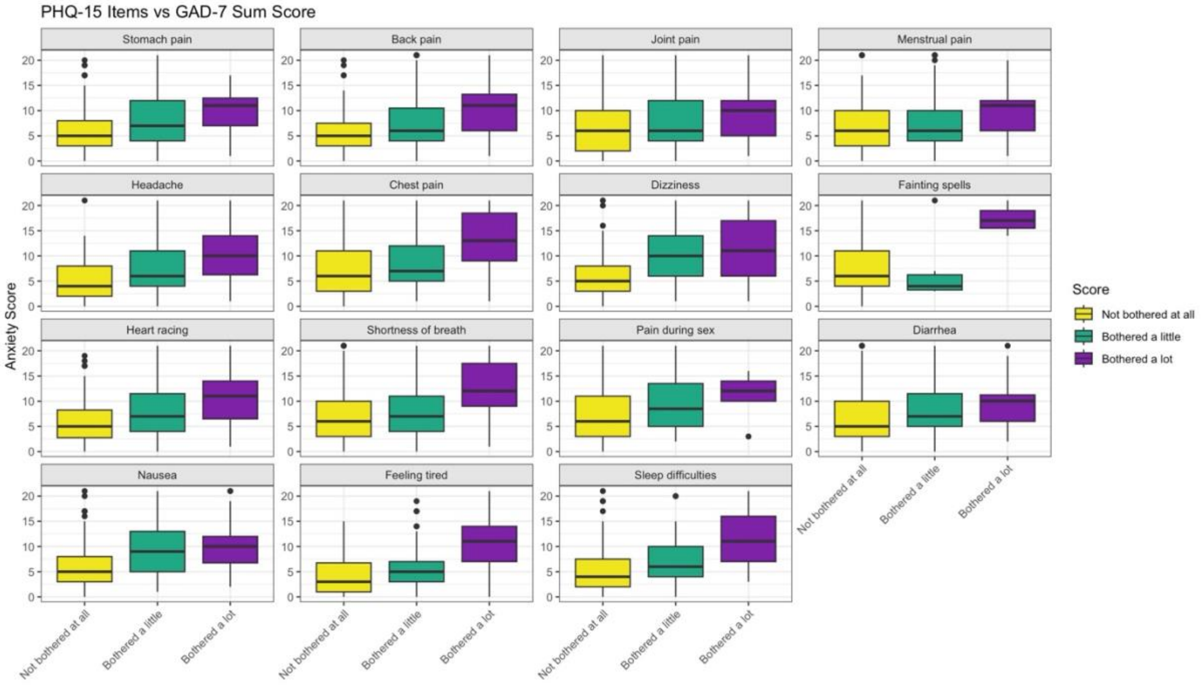
PHQ-15 Items vs GAD-7 Sum Score
Stomach pain
Back pain
Joint pain
Menstrual pain
Headache
Chest pain
Dizziness
Fainting spells
Heart racing
Shortness of breath
Pain during sex
Diarrhea
Nausea
Feeling tired
Sleep difficulties
Anxiety Score
Score
Not bothered at all
Bothered a little
Bothered a lot

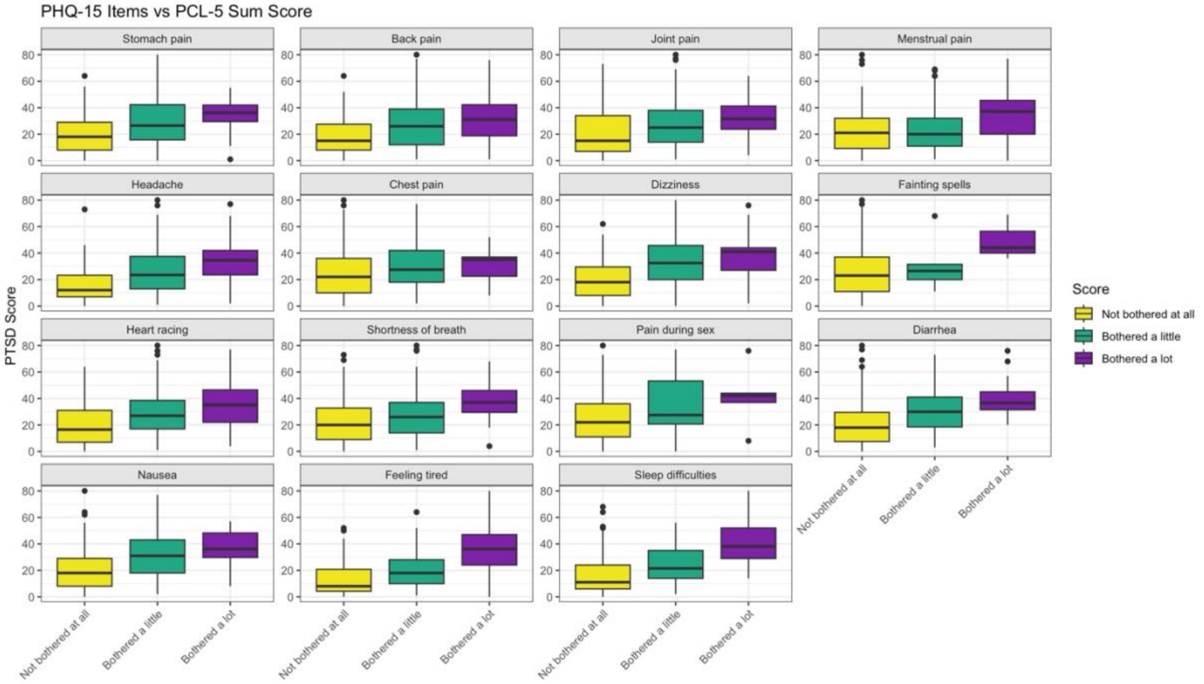
PHQ-15 Items vs PCL-5 Sum Score
Stomach pain
Back pain
Joint pain
Menstrual pain
Headache
Chest pain
Dizziness
Fainting spells
Heart racing
Shortness of breath
Pain during sex
Diarrhea
Nausea
Feeling tired
Sleep difficulties
PTSD Score
Score
Not bothered at all
Bothered a little
Bothered a lot

**Figure S11**. Boxplots of PHQ-9, GAD-7 and PCL-5, stratified by somatic symptom severity for MAP-U (SE).

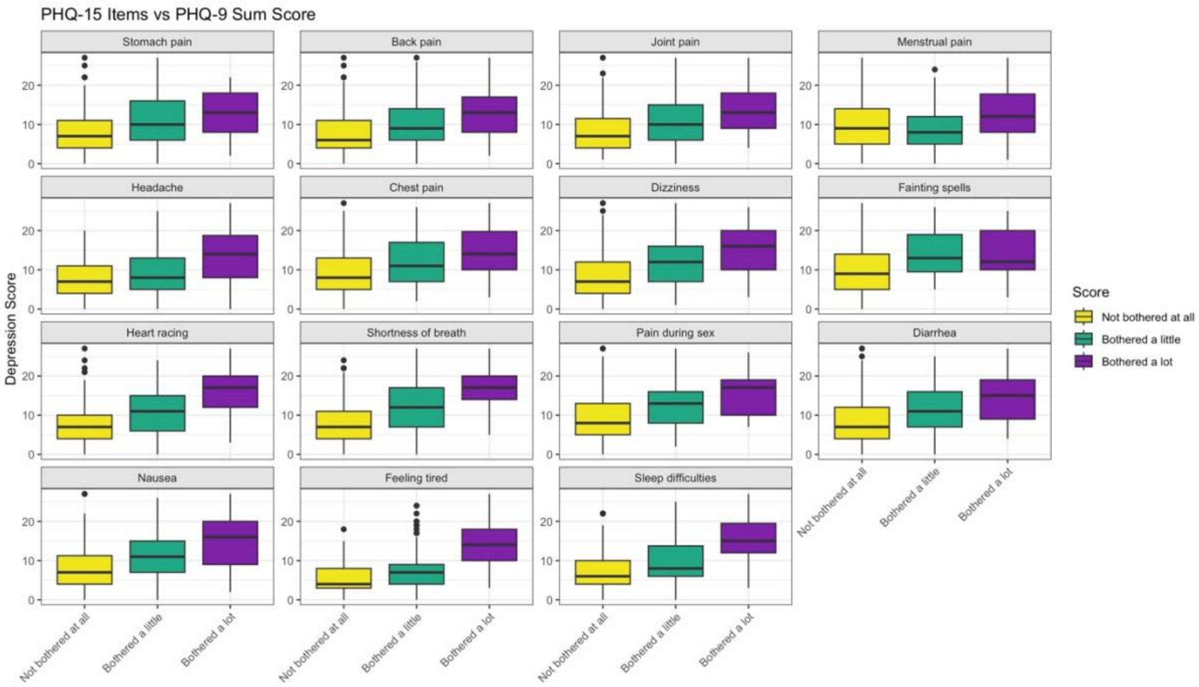

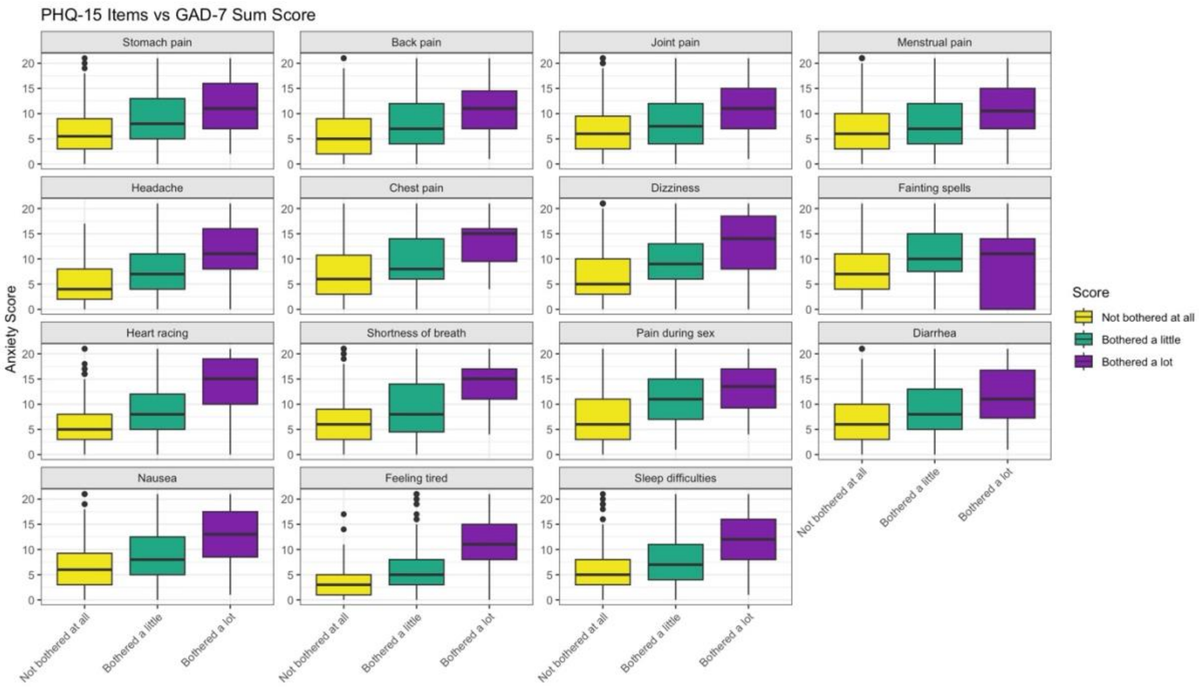

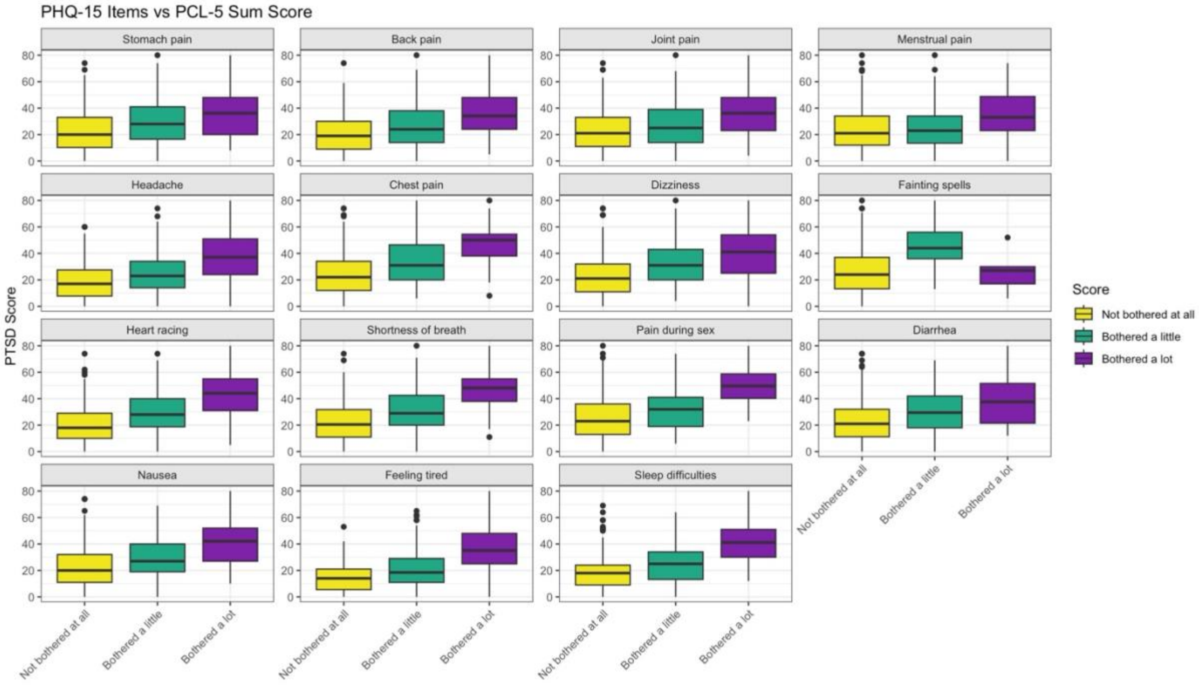
